# Exploring reward learning disruptions as a possible mechanism underlying neuropsychiatric symptoms in Parkinson’s disease

**DOI:** 10.64898/2026.01.14.26344132

**Authors:** Sophie Sun, Madeleine Sharp

**Author notes:** Corresponding Author Sophie Sun, 251-3801 rue University Montreal, QC, H3A 2B4.

## Abstract

**Background:** The causes of neuropsychiatric symptoms in Parkinson’s disease (PD) remain ill-defined. Disruptions in dopamine-dependent reward learning, a consequence of midbrain dopamine loss, potentially represent a mechanism that could underlie the neuropsychiatric symptoms of apathy, depression, and impulsivity, all of which have been proposed to reflect aberrant goal-directed behaviour. However no large-scale investigation that jointly considers these symptoms in PD has ever been undertaken. We aimed to determine if reward learning is associated with apathy, depression and impulsivity symptom in two samples of PD patients.

**Methods:** Two samples of PD patients (n_sample1_=81, n_sample2_=90), tested in their medicated state, completed two widely used reward learning tasks (probabilistic stimulus selection task, probabilistic reward task) from which we derived five summary measures of performance. Apathy, depression, and impulsivity were evaluated using validated self-report questionnaires.

**Results:** Overall, there was poor consistency in the relationship between reward learning and neuropsychiatric symptom severity. Greater depressive symptom severity was associated with slower reward learning performance in sample 1, but with both slower and faster reward learning performance in sample 2. Greater impulsivity symptom severity was associated with slower reward learning performance in sample 1 but not in sample 2. There were no associations between apathy and reward learning.

**Conclusions:** We found inconsistent relationships between symptoms of apathy, depression, and impulsivity and reward learning performance across two samples of PD patients. While this doesn’t rule out the possibility that reward learning impairments contribute to these symptoms, it suggests any effect is likely to be small and overshadowed by other non-measured factors.

## Introduction

Neuropsychiatric symptoms are prominent in Parkinson’s disease (PD), and a striking yet unresolved aspect of these symptoms is the degree to which they vary across individuals^1–4^. Early neurodegeneration in PD involves several brainstem nuclei responsible for ascending neuromodulatory systems – including the dopaminergic, serotonergic, and noradrenergic pathways – that are thought to contribute to the emergence of neuropsychiatric symptoms^4–6^. Of these, dysfunction in the dopaminergic system is a particularly promising candidate mechanism given its well-established role in reward learning^7,8^. In psychiatric populations, reward learning has been linked to goal-directed mood and behavioural symptoms such as apathy, depression and impulsivity, symptoms that are also common in PD^9–13^. Reward learning impairments have been consistently observed in PD^14–16^ and a recent study showed that the severity of degeneration in the dopaminergic substantia nigra is associated with reward learning performance in PD^17^. Together, these observations raise the possibility that reward learning could represent a shared mechanism across goal-directed neuropsychiatric symptoms in PD.

There is some indication from recent research that a relationship between reward learning performance and neuropsychiatric symptoms exists in PD, but most studies have focused on examining symptoms in isolation of one another, precluding the identification of mechanisms that can generalize across symptoms that manifest as altered goal-directed behaviour. For instance, it has been shown that reward learning performance is impaired in PD patients with depression^18–20^ and in patients with apathy^21,22^. Impulse control disorders (ICDs), which typically manifest in relation to dopaminergic treatment, have also been associated with alterations in reward learning but in the opposite direction: PD patients exhibit a hypersensitivity to reward manifesting behaviourally as faster value updating in response to feedback, i.e. a faster reward learning rate^23,24^. A meta-analysis of 55 PD studies where various measures of reward learning were collected also suggested that reward processing impairments are a promising causal mechanism for neuropsychiatric symptoms in PD. Notably, none of these studies measured more than one mood symptom, none included more than one reward learning measure, and all were conducted on relatively small samples (median sample size across studies was 24). As a result of these limitations, the authors concluded that it was not possible to make meaningful conclusions based on the existing evidence^25^.

The idea that mood symptoms could share a common underlying disruption in goal-directed behaviour is consistent with the notion in psychiatry that different psychiatric disorders, primarily defined based on their behavioural manifestations, may in fact share common underlying neural and cognitive mechanisms. The Research Domain Criteria (RDoC) framework^26^ outlines various behavioural domains that help operationalize possible mechanisms including a positive valance system encompassing reward learning performance. Given that the neural correlates of reward learning are relatively well-established, identifying reward learning disruptions as a common feature of apathetic, depressive, and impulsive symptoms in PD would provide promising targets for interventions and proximal measurements to track effectiveness.

In the present study, we aimed to investigate whether individual differences in reward learning performance are associated with symptoms of apathy, depression, and impulsivity in PD. We hypothesized that slower reward learning rates would be associated with higher apathy and depression symptom severity, and that faster reward learning rates would be associated with higher impulsivity symptom severity. We examined this hypothesis in two large independent cohorts of PD patients who were evaluated in their usual dopaminergic medication state and completed self-report mood questionnaires and two RDoC-recommended reward learning tasks (the probabilistic stimulus selection task^27^ and the probabilistic reward task^28^). Importantly, performance on these tasks has been associated with at least one of the following: dopamine transmission, activation of dopamine-associated brain regions, and degeneration of the dopaminergic substantia nigra^15,17,27–33^. Since many different outcome measures of reward learning have previously been extracted from these tasks in relation to PD, to dopamine state, and to mood in non-PD populations, we chose to jointly examine five possible outcome measures of performance.

## Methods and Materials

### Participants in Sample 1

Eighty-one people with Parkinson’s disease (PD) were included in the present study as a discovery sample. These participants were sourced from a larger in-person cohort of participants who were recruited from the Quebec Parkinson Network^34^. The only exclusion criterium was age below 50 years old to ensure a representative sample. Participants were asked to complete the study in their usual medicated state (i.e., on dopaminergic medication) in their language of preference (English/French), provided written informed consent, and were compensated $25. The study protocol was approved by the McGill University Health Centre Research Ethics Board.

### Participants in Sample 2

A total of 90 people with PD were included as a replication sample. These participants were sourced from a larger online cohort of participants who were recruited from the Canadian Open Parkinson Network. The only exclusion criterium was age below 50 years old to ensure a representative sample. Participants were asked to complete the study in their usual medicated state and in either English or French. Participants were entered to win one of ten $100 Amazon e-gift cards. The study was completed entirely online and was approved by the McGill University Health Centre Research Ethics Board.

Due to the online and unsupervised nature of the study, not all participants completed all elements of the protocol. For the purposes of replicating our analyses, we included participants who had completed the probabilistic stimulus selection task and at least one of the neuropsychiatric symptom questionnaires of interest. The relevant sample sizes for each combination of task and questionnaire are included in **Figure 3**.

### Reward Learning Tasks

*Probabilistic Stimulus Selection Task.* Reward learning performance was assessed using an adapted probabilistic stimulus selection task that has been widely administered in PD patients^17,27,35^. The task has two parts: a training phase and a test phase.

In the training phase of the task, participants had to learn from trial-and-error about the different reward values of three pairs of stimuli over 150 trials (**Supplementary Fig. 1A**). The possible reward probabilities for each pair were 0.8 and 0.2 (AB pairing), 0.7 and 0.3 (CD pairing), and 0.6 and 0.4 (EF pairing). Full details of task administration were recently described (Sun et al., 2025).

In the test phase of the task, participants were shown the same three training pairs (i.e., AB, CD, EF) and eight new combinations of the stimuli, where the most likely rewarded stimulus (A) the least likely rewarded stimulus (B) are each paired with all other stimuli over 110 trials (AC, AD, AE, AF, BC, BD, BE, BF; **Supplementary Fig. 1B**). Participants were tasked to select the stimulus associated with greater probability of reward to measure if reward values from the training phase were learned and generalize to new contexts. Further details in the Supplementary Materials.

*Probabilistic Reward Task.* We also used a probabilistic reward task that has been previously administered in psychiatric populations as a secondary measure of reward learning^28,32,33,36–38^. Conceptually, the task aims to implicitly bias participant responses by rewarding correct responses to one stimulus three times more often than correct responses to the other stimulus. Further details are included the in Supplementary Materials.

### Self-report neuropsychiatric symptom questionnaires

Self-report questionnaires were completed online for all participants, including sample 1, where this was done to minimize the length of the in-person visit. The battery consisted of questionnaires for symptoms of depression (Geriatric Depressive Scale (GDS-15), short form ranging from 0-15 points^39–41^), anhedonia (Snaith Hamilton Pleasures Scale (SHaPS), ranging from 0-42 points^42^), apathy (Apathy Evaluation Scale (AES), ranging from 18-72 points^43^), and impulsivity (Barratt Impulsiveness Scale version 11 (BIS-11), ranging from 30-120 points^44^; Questionnaire for Impulsive-Compulsive Disorders in Parkinson’s Disease (QUIP), ranging from 0-112 points^45^).

### Analysis

To assess our cross-sample generalizability, sample differences in demographic and clinical characteristics and neuropsychiatric symptom scores were evaluated using independent samples t-tests, Mann-Whitney U tests, and chi-squared tests.

Our behaviour of interest was reward learning. Existing studies of reward learning have examined reward learning using several different measures, so we computed five different outcome measures from performance on the reward learning tasks: positive learning rate, learning slope, and approach ‘A’ accuracy were estimated from the probabilistic stimulus selection task, while response bias and rich-lean hit rate were estimated from the probabilistic reward task (**Table 1**). We also evaluated the sample differences for each outcome measure, which is described in greater detail in the supplementary materials.

**Table 1.**
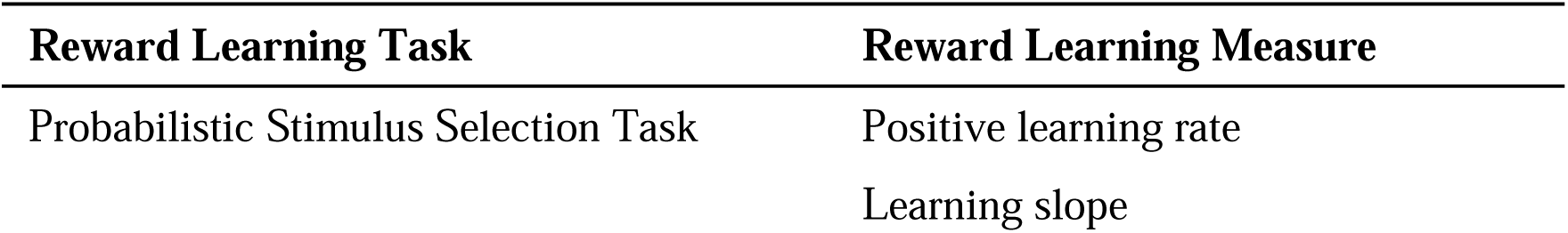
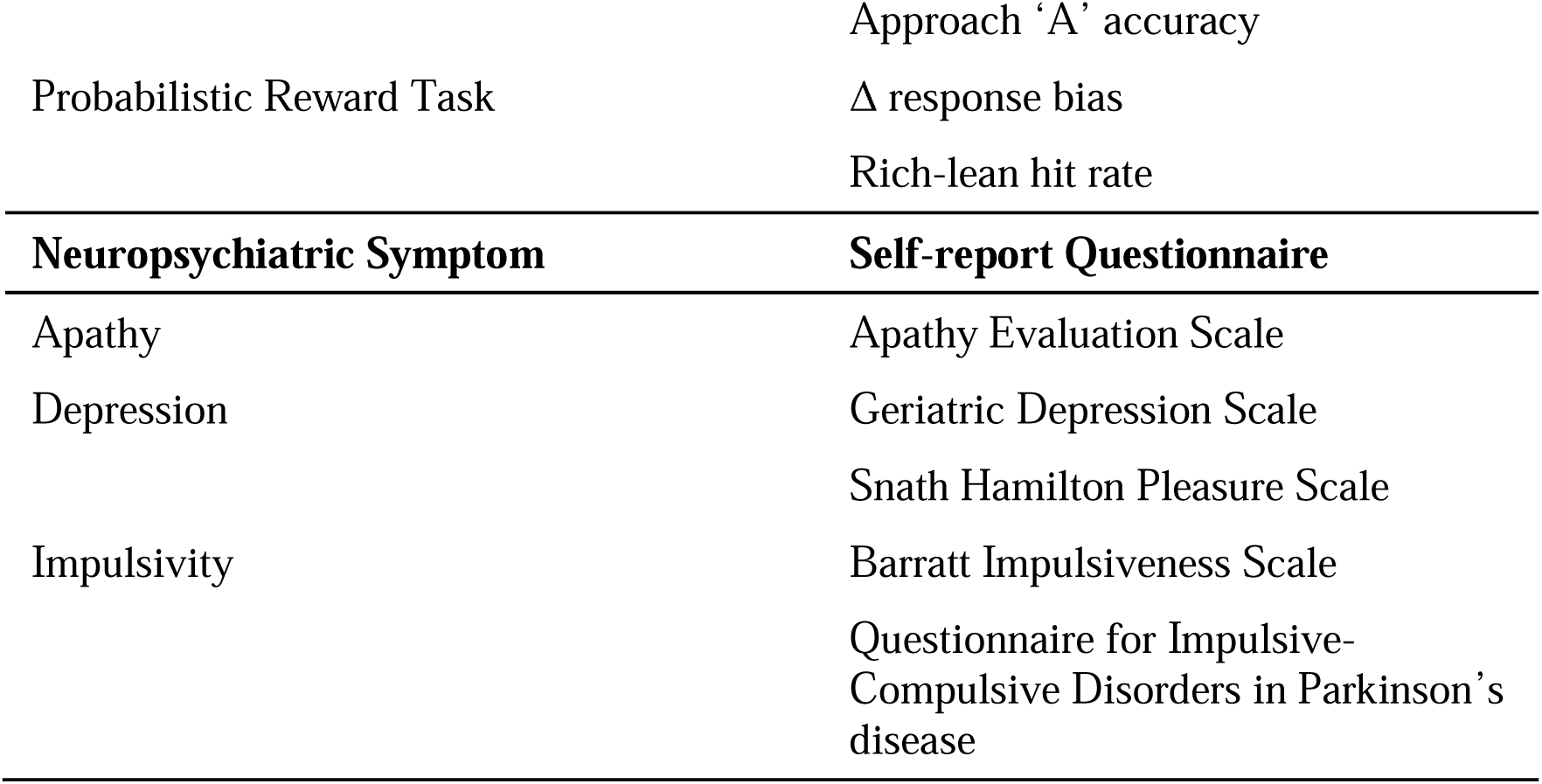
Summary of Measures.

To examine the relationships between reward learning performance and neuropsychiatric symptom severity, we performed 1) multiple linear regressions using each reward learning summary measure as the independent variable and each symptom score as the dependent variable in separate analyses in each sample and 2) a canonical correlation analysis (CCA) including all five summary measures of reward learning and all five neuropsychiatric symptom questionnaire total scores as variable sets collapsed across both samples. Age, disease duration, and sex were included as covariates in all regressions. Sex was effect coded (Males as the reference group). All continuous variables were z-scored. Statistical significance was defined as p < 0.05. Regression p-values were False Discovery Rate (FDR)-corrected to account for multiple comparisons. A CCA p-value was obtained by permutation testing (n=5000).

Lastly, we performed exploratory analyses in sample 1 investigating 1) the Pearson correlations between the five measures of reward learning performance and levodopa equivalent dose and 2) differences in impulsivity scores (QUIP and BIS-11) across medication regimens (regimens: levodopa without dopamine agonist and levodopa with dopamine agonist) using a one-way ANOVA.

## Results

### Sample Characteristics

On average, sample 1 was younger and had lower disease duration compared to sample 2 (**Table 2**). The difference in both age and disease duration was about 3 years. Age and disease duration were not correlated (R_sample1_=0.10, p=0.40; R_sample2_=0.002, p=0.98), so to ensure that these sample differences did not affect our analyses, we included both variables as covariates in all analyses. Otherwise, there were no significant differences between samples including in our variables of interest, i.e., no sample differences across the neuropsychiatric symptoms scores (*p*s>0.20; **Fig 1**) nor across the reward learning measures (*p*s>0.11; **Fig 2**). The neuropsychiatric symptom scores had small to medium correlations with each other across both samples (**Supplementary Figure 2A-B).** No PD medication dose information was collected in Sample 2.

**Figure 1.**
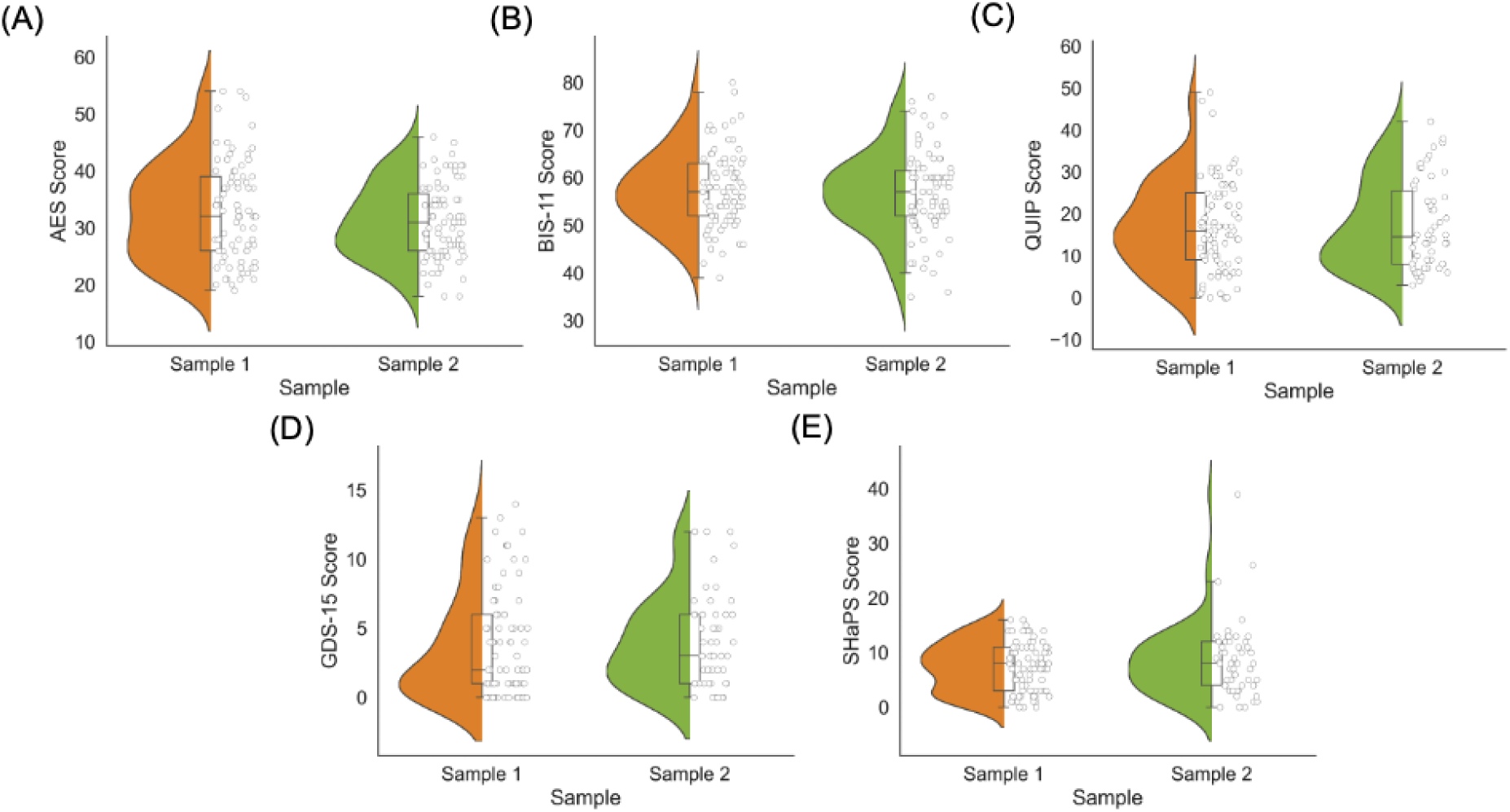
Neuropsychiatric symptom scores across samples. Samples do not differ across A) Apathy Evaluation Scale (AES) scores (p = 0.32; possible scores range from 18-72), B) Barratt Impulsiveness Scale version 11 (BIS-11) scores (p = 0.54; possible scores range from 30-120), B) Questionnaire for Impulsive-Compulsive Disorders in Parkinson’s disease (QUIP) scores (p = 0.46; possible scores range from 0-112), D) Geriatric Depression Scale short form (GDS-15) scores (p = 0.21; possible scores range from 0-15), or E) Snaith Hamilton Pleasure Scale (SHaPS) scores (p = 0.75; possible scores range from 0-42).

**Figure 2.**
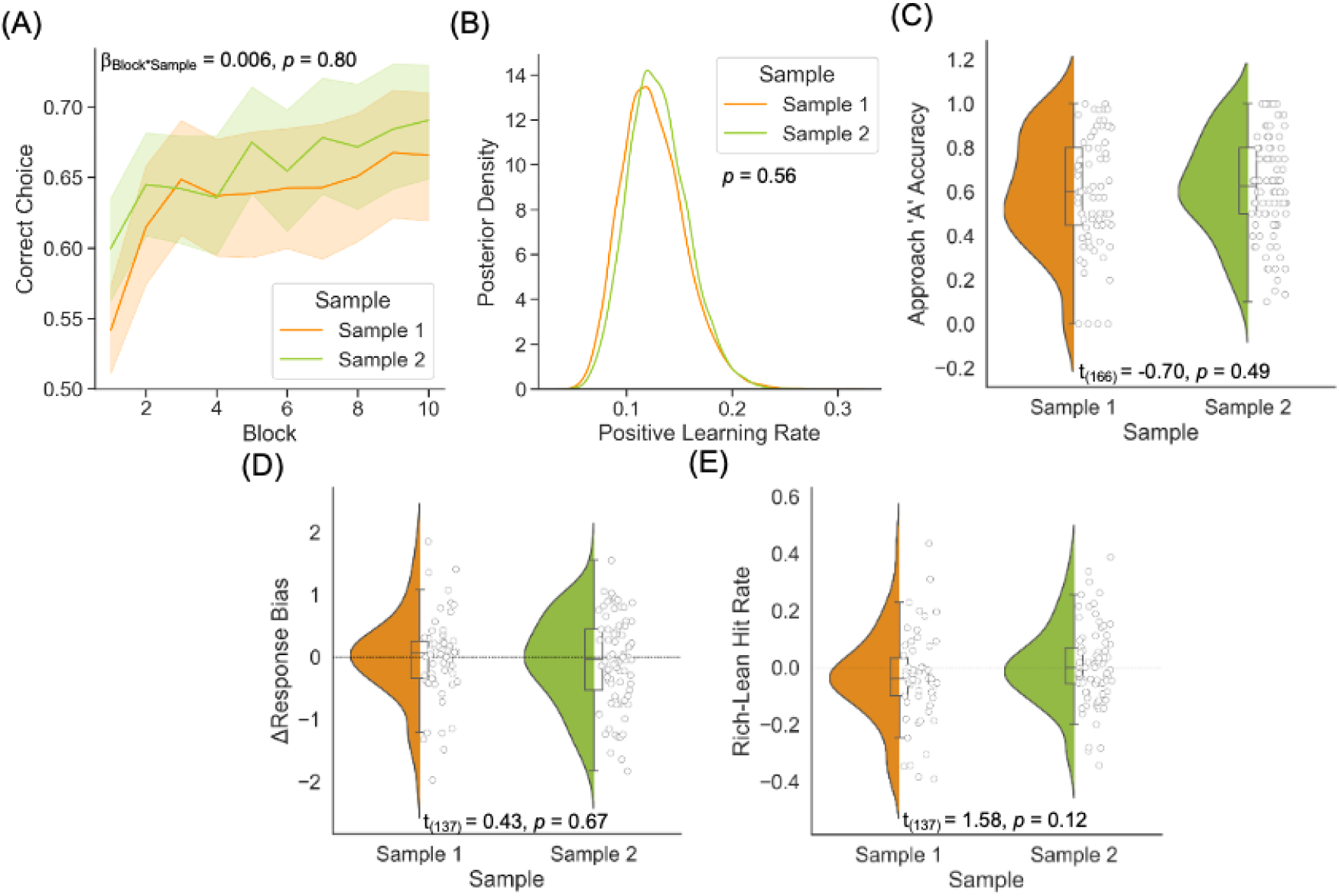
Reward learning performance across samples. Samples did not differ in (A) learning slopes, (B) positive learning rates, (C) approach ‘A’ accuracy, (D) Δ response bias (block 3-block1) and (E) the difference in hit rates between rich and lean stimuli (rich-lean).

**Table 2.** Sample Characteristics.

|  | **Sample 1**<br>**(n=81)** | **Sample 2**<br>**(n=90)** | **p-value** |
| --- | --- | --- | --- |
| Age | 64.2 (7.9) | 67.2 (7.4) | **0.01<sup>a</sup>** |
| Males | 56 | 57 | 0.52<sup>b</sup> |
| Education, years | 15.2 (3.1) | 15.6 (3.4) | 0.45<sup>a</sup> |
| Disease Duration | 4.3 (3.7) | 7.2 (4.9) | **<0.001<sup>c</sup>** |
| LED (mg) | 827.2 (597.0)<sup>d</sup> |  |  |
| # also on DA agonist | 15 |  |  |
| # also on MAO-B inhibitor | 14 |  |  |
| # also on COMT inhibitor | 13 |  |  |
| # not on PD medication | 2 |  |  |

Values depict mean and standard deviation in brackets.
Abbreviations: Levodopa Equivalent Dose (LED)
<sup>a</sup>Group comparison evaluated using parametric independent samples t-test
<sup>b</sup>Group comparison evaluated using chi-square test
<sup>c</sup>Group comparison evaluated using non-parametric Mann-Whitney U test
<sup>d</sup>Medication information was only available for 62/81 PD patients

### Relationships between reward learning and each neuropsychiatric symptom

In sample 1, we found variable relationships between the different reward learning performance measures and neuropsychiatric symptom scores. First, there was a negative effect of positive learning rate on QUIP and GDS-15 scores in that lower positive learning rate was associated with greater symptom score, but only the effect on QUIP score survived FDR-correction (QUIP: *β _positive learning rate_*= -0.32, *p*=0.003, *p_FDR_*=0.02; GDS-15: *β _positive learning rate_*= -0.29, *p*=0.01, *p_FDR_*=0.07; **Fig. 3B**). There was no effect of positive learning rate on AES, BIS-11, and SHaPS scores (*p*s>0.05, full model results in **Supplementary Table 2**).

**Figure 3.**
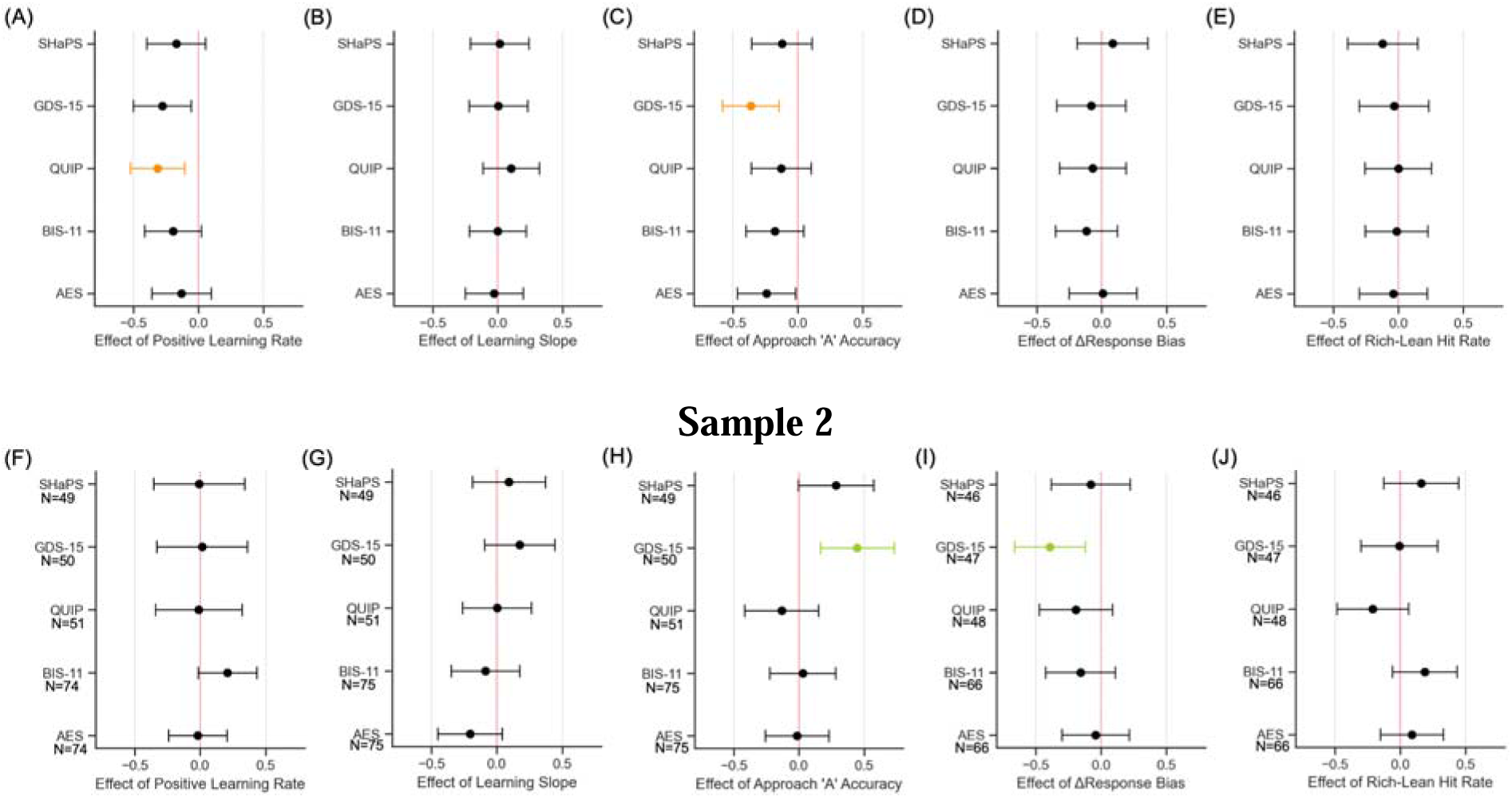
Effect of reward learning summary measures on neuropsychiatric symptom scores. There were no effects of learning slope on any neuropsychiatric symptom score in both the sample 1 (A, n=80) and sample 2 (F). There were negative effects of positive learning rate on impulsivity (QUIP) and depression (GDS-15) symptom scores (B, n=79), but this was not reproduced (G). There were negative effects of approach ‘A’ accuracy on depression apathy (AES) and depression (GDS-15) symptom scores (C, n=77), but this was not reproduced (H). There were no effects of Δ response bias on any neuropsychiatric symptom score in the sample 1 (D, n=59). However, there was a negative effect of Δ response bias on depression (GDS-15) symptom score in sample 2 (I). There were no effects of rich-lean hit rate on any neuropsychiatric symptom score in both sample 1 (E, n=59) and sample 2 (J). Points represent the standardized beta estimates, allowing comparisons of beta estimates across models, and error bars represent the 95% confidence interval. Coloured points and error bars represent effects that survived False Discovery Rate-correction.

Next, there was no effect of learning slope on any neuropsychiatric symptom score (*p*s>0.61, **Fig. 3A**, full model results in **Supplementary Table 3**).

There was an expected negative effect of approach ‘A’ accuracy on AES and GDS-15 score where lower approach ‘A’ accuracy was associated with greater symptom score, but only the effect on GDS-15 score survived FDR-correction (AES: *β _approach ‘A’ accuracy_*=-0.23, *p*=0.05, *p_FDR_*=0.10; GDS-15: *β _approach ‘A’ accuracy_*=-0.32, *p*=0.01, *p_FDR_*=0.01; **Fig. 3C**). There was no effect of approach ‘A’ accuracy on BIS-11, QUIP, or SHaPS scores (*p*s>0.17, full model results in Supplementary Table 4).

Lastly, there were no effects of Δ response bias nor of rich-lean hit rate on any neuropsychiatric symptom score (*p*s>0.37, **Fig. 3D** and **Fig. 3E** respectively, full model results in **Supplementary Table 5** and **Supplementary Table 6**).

### Replication of individual relationships between reward learning and neuropsychiatric symptoms

Overall, the statistically significant relationships identified in sample 1 were not reproduced in sample 2. First, there were no associations between positive learning rate and any of the neuropsychiatric symptom scores (*p*s>0.07, **Fig 3F**, full model results in **Supplementary Table 7**). There were also no associations between learning slope and any of the neuropsychiatric symptom scores (*p*s>0.06, **Fig 3G**, full model results in **Supplementary Table 8**). Furthermore, we found an unexpected positive effect of approach ‘A’ accuracy on GDS-15 score where greater approach ‘A’ accuracy was associated with greater depression score, which survived FDR-correction (**Fig 3H**, GDS-15: *β _approach ‘A’ accuracy_*=0.45, *p*=0.003, *p_FDR_*=0.01), but this relationship is in the opposite direction as the relationship found in sample 1. Otherwise, there were no associations between approach ‘A’ accuracy and AES, BIS-11, QUIP, and SHaPS scores (*p*s > 0.05, full model results in **Supplementary Table 9**).

From the probabilistic reward task, we found no association between Δ response bias and AES, BIS-11, QUIP, and SHaPS scores (**Fig 3I**, *p*s>0.17, full model results in **Supplementary Table 10**). However, there was a new negative effect of Δ response bias on GDS-15 score where a smaller Δ response bias was associated with greater depression score and this effect survived FDR-correction (GDS-15: *β_Δ response bias_*=-0.39, *p*=0.01, *p_FDR_*=0.03). Lastly, there was no association between rich-lean hit rate and any of the neuropsychiatric symptom scores (**Fig 3J**, *p*s>0.26, full model results in **Supplementary Table 11**).

### Exploratory medication analyses

We performed an exploratory analysis in participants in sample 1 with available PD medication information (n=62) to probe the possible influence of dopamine medication on reward learning performance. While there were weak negative relationships between daily levodopa equivalent dose and reward learning performance on three of five outcome measures, we did not find any significant correlations (ps>0.10; **Supplementary Figure 3**).

Since the development of ICDs in PD have been closely related to the administration of dopamine agonists, we explored whether impulsivity scores differed in PD patients who were taking levodopa without dopamine agonists (n=43) and with dopamine agonists (n=15). We found that QUIP and BIS-11 scores were on average greater in those taking dopamine agonists compared to those who were not, but this difference was not statistically significant (*t*=1.98, *p*=0.05; *t*=0.94, *p*=0.35; **Supplementary Figure 4**). Together, these exploratory analyses suggest that dopaminergic medication likely had minimal influence on our findings in sample 1.

### Correlation between reward learning summary measures

Given the inconsistent relationships between reward learning and neuropsychiatric symptom severity observed across task measures both within and between samples, we examined the correlations across reward learning summary measures (**Supplementary Figure 2C-D**). Th correlation matrices for both samples demonstrated that the five reward learning summary measures were mostly uncorrelated (absolute r between 0.02-0.35).

### Correlation between reward learning performance and neuropsychiatric symptoms

Since we aimed to investigate whether disruptions in reward learning performance are an underlying mechanism common to apathy, depression, and impulsivity symptoms in PD, we tested this relationship using a multivariate approach where all reward learning measures and all neuropsychiatric symptom scores were entered into a CCA collapsed across both samples. Results revealed a moderate relationship between reward learning performance and goal-directed neuropsychiatric symptoms in PD patients, but this was not a significant correlation (*r*=0.30, *p*=0.92, **Figure 4**).

**Figure 4.**
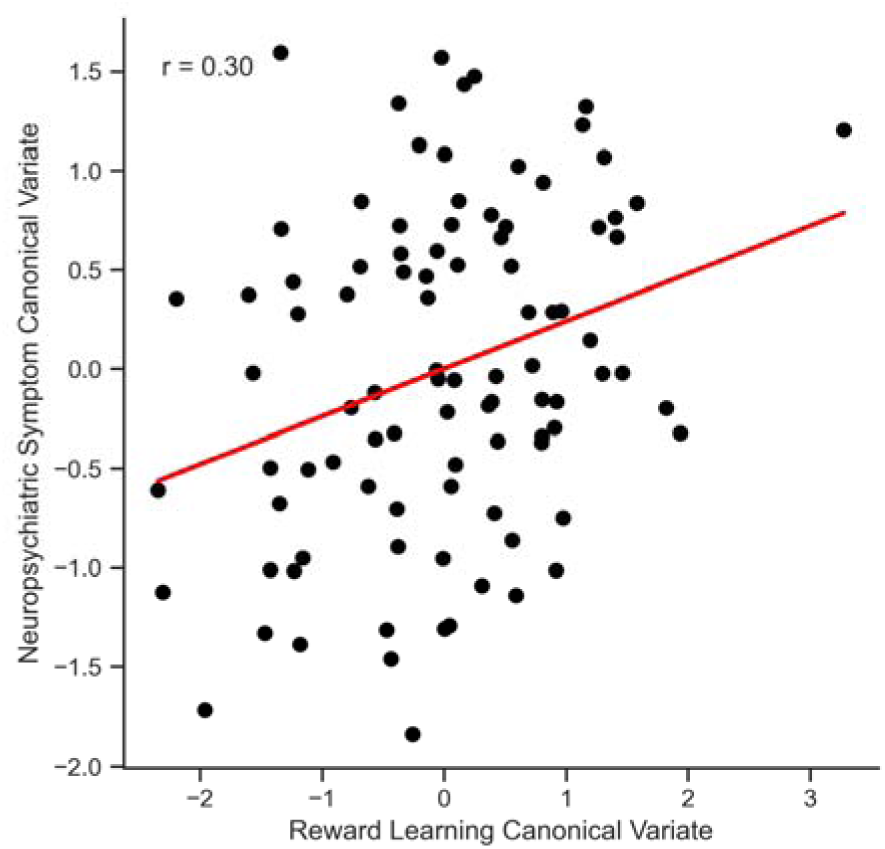
Correlation between reward learning performance and neuropsychiatric symptoms as canonical variates. The reward learning canonical variate is comprised of positive learning rate, learning slope, approach ‘A’ accuracy, Δ response bias, and rich-lean hit rate summary measures. The neuropsychiatric symptom canonical variate is comprised of scores on the AES, BIS-11, QUIP, GDS-15 and SHAPS. The correlation between the reward learning and neuropsychiatric canonical variates was not significant (*r*=0.30, *p*=0.92).

## Discussion

In the present study, we examined whether reward learning performance was associated with neuropsychiatric symptoms in two independent samples of PD patients. Overall, we found surprisingly inconsistent relationships between reward learning performance and symptoms of apathy, depression, and impulsivity across both samples. This suggests that while individual differences in reward learning disruptions in PD *may* contribute to neuropsychiatric symptom severity, there may be other factors that attenuate this relationship and other candidate mechanisms that more stably predict these neuropsychiatric symptoms in PD.

First, we found some evidence of a relationship between reward learning performance and depressive symptoms: worse approach ‘A’ accuracy and Δ response bias (indicating worse reward learning) were associated with greater depression severity in samples 1 and 2, respectively. This is consistent with previous studies demonstrating that reward learning performance is impaired in both PD and non-PD patients with depression compared to controls^10,18–20^. However, in sample 2, there was also an unexpected relationship in the opposite direction: better approach ‘A’ accuracy (i.e., better reward learning) was associated with higher depression severity. While puzzling, it is worth noting that the depression scores in sample 2 were more weakly correlated with the other neuropsychiatric symptoms than in sample 1, suggesting that sample 2 might have had a different expression of depressive symptoms. Overall, these findings suggest that the association between reward learning and depression in PD is present but modest and may be sensitive to sample composition and measurement context. The fact that we observed effects in opposite directions across samples argues against a simple, robust linear relationship and instead suggests that depression-related variance in reward learning performance is easily overshadowed by other sources of heterogeneity (e.g., medication status, comorbid symptoms, fluctuating psychosocial factors).

In the case of apathy, we did not find a relationship with reward learning performance. This opposes previous studies reporting a relationship between reward learning impairments, measured across various paradigms, and apathy in PD patients^13,21,22,47,48^. However, it should be noted that these past studies focused on clinically apathetic PD patients using either formal diagnoses or cutoff scores on apathy scales, whereas our recruitment strategy was intentionally inclusive to ensure a broad and representative sample and did not specifically focus on clinical subgroups. As a result, apathy levels in our samples were lower (sample 1: 30%, sample 2: 17%, using cutoff score of 38 on the AES^48^) compared to depression levels (sample 1: 35%, sample 2: 23%, using cutoff score of 5 on the GDS-15), possibly reflecting the fact that the presence of apathy could hinder research participation. Since prior literature focused on clinically apathetic patients, our sampling of a wide range of apathy levels may have diluted the expected strength of the relationship between reward learning performance and apathy symptom severity.

Lastly, we found a negative relationship between reward learning performance and impulsivity in sample 1 such that lower positive learning rate (i.e., worse learning from reward) was associated with greater symptoms of impulsivity/compulsivity, as measured by the QUIP. This finding was also unexpected given previous studies demonstrating that sensitivity to reward, in particular the rate at which expectations are updated by rewarding feedback, is exaggerated in PD patients with ICD^23,49–51^, but this literature has also yielded mixed findings^4,25^. Dopamine medications in PD, specifically dopamine receptor agonists, are thought to overstimulate learning from rewards, especially in patients with ICDs, and dampen learning from losses^23,27,29^. However, in an exploratory analysis relying on the small subsample of individuals taking a dopamine agonist, we found no relationship between dopamine receptor agonist use and impulsivity symptom severity. We also found no significant associations between levodopa equivalent daily dose and impulsivity symptom severity. Together, this could suggest that our observation that slower reward learning rate was associated with higher impulsivity may reflect the presence of an additional cognitive deficit caused by PD, such as loss of cognitive control, that we did not account for but that could also influence the manifestation of impulsivity.

Finally, we hypothesized that reward learning disruptions were a common mechanism underlying apathy, depression, and impulsivity in PD, so we conducted a multivariate correlation between aggregated reward learning and aggregated neuropsychiatric symptom measures. A CCA revealed a weak but not statistically significant relationship between the reward learning canonical variate (composed of the five reward learning performance measures) and the neuropsychiatric symptom canonical variate (composed of the five neuropsychiatric symptom scores). Taken with the overall inconsistent univariate relationships between reward learning performance and symptoms of apathy, depression, and impulsivity across both PD samples, this suggests that if reward learning disruptions are a common mechanism of these goal-directed neuropsychiatric symptoms in PD, the effect is small and several other factors may be affecting the robustness of this effect including sampling, construct validity of the reward learning measures, and the presence of individual differences occurring due to unmeasured factors.

The sampling for this study included two cohorts of individuals with PD patients who completed all assessments on their regular dopamine regimens, which may have attenuated expected relationships given that dopamine medication has been shown to remediate reward learning deficits^14,51–54^, though this would not have been expected to negatively influence the relationship to impulsivity. The decision to test individuals on medication was made for both practical and conceptual reasons. First, we aimed for large-scale testing and to recruit a more representative sample, the feasibility of which would have been hindered by requiring participants to withdraw from their medications for 12-16 hours. Second, neuropsychiatric symptoms exist even in PD patients who experience good control of motor symptoms and minimal-to-no OFF time (though in some individuals, mood symptoms are exacerbated when OFF)^55,56^. This suggests that the mechanism we are investigating is partially but not entirely remediated by dopaminergic medications. We therefore reasoned that testing patients on medications would facilitate recruitment while nonetheless capturing the symptom experiences of interest. However, the potentially important role of medication-related mood fluctuations should be considered in future research aiming to identify underlying mechanisms, especially considering evidence that degeneration in the dopaminergic system (and therefore potentially modulation of this system) influences other neurotransmitter systems^2^.

There is also the potential issue of construct validity for tasks measuring reward learning behaviour. In the present study, we examined reward learning behaviour using five outcome measures extracted from two commonly used tasks. We specifically chose tasks and measures that have been previously associated with dopamine transmission, activation of dopamine-associated brain regions, and/or degeneration of the dopaminergic substantia nigra^15,17,27–33^. Yet, these five measures were only weakly, and not significantly, correlated with one another. Assuming each of these measures is measuring a meaningful behaviour, then it suggests they may be indexing different processes rather than a reward learning latent construct. However, the lack of correlation across measures likely also points to the increasingly recognized challenge of using cognitive task performance as a reliable measure of individual differences. Indeed, some recent work has demonstrated poor to moderate reliability of reward learning measures and that reward learning tasks yield greater within subject variance than self-report survey-based methods^57–59^. These concerns were also previously raised in studies of self-regulation tasks, indicating that further efforts are required to develop and implement reliable task-based cognitive assessments^60,61^.

Furthermore, individual differences related to unmeasured consequences of PD on the brain, as well as other psychological and cognitive factors, may also be playing a role. For instance, it is likely that degeneration in other neurotransmitter systems also contributes^2^. Serotonergic degeneration is thought to play a role in mood symptoms of PD, particularly depression, with one PET study showing that PD patients with depression exhibit altered serotonergic binding compared to patients without depression^62^ and another study finding that patients with depression have lower serotonin levels in the cerebrospinal fluid^63^. Noradrenergic and cholinergic systems, which support cognitive processes like executive function and attention and are also impaired in PD^64–66^, may also contribute to certain neuropsychiatric symptoms. Future studies should jointly consider the contributions and potential interactions of degeneration in these neurotransmitter systems.

In summary, we did not find robust evidence that individual differences in reward learning are strongly associated with neuropsychiatric symptoms in PD, whether indexed by summary measures of raw task performance or by computationally derived parameters. The few significant associations we observed, mainly for depressive symptoms, were small and inconsistent across samples, tasks, and measures. These findings do not rule out a true relationship, but suggest that any effect of reward learning on neuropsychiatric symptoms is likely modest and not the dominant behavioural mechanism in PD. More generally, our results underscore the difficulty of investigating cognitive mechanisms of neuropsychiatric symptoms in PD – a knowledge gap that remains critical. Future work will have to carefully consider the choice and optimization of task-based measures of an underlying cognitive process of interest, including temporal dynamics of symptoms. For instance, PD-related changes in reward learning may exert an effect on mood (e.g., causing depressive symptoms), but within person variability in both learning performance and mood may obscure such links. Nonetheless, because PD involves relatively well-defined and increasingly measurable regional and circuit pathology, it provides a valuable model system in which to interrogate the mechanisms of neuropsychiatric symptoms with findings that could generalize to other disorders.

## Data Availability

All data produced in the present study are available upon reasonable request to the authors

## Acknowledgments

Yilin Zhang, Nasri Balit, Sarah Bogard, Roozbeh Sattari, Courtney Jackson, Lucy Zou, QPN, COPN, and all participants.

This research was supported by a grant from the Canadian Institutes of Health Research (MS), a salary award from the Fonds de Recherche du Quebec – Santé, and a graduate student award from Parkinson Canada (SS).

## References

1. Weintraub D, Aarsland D, Chaudhuri KR, Dobkin RD, Leentjens AF, Rodriguez-Violante M, et al. The neuropsychiatry of Parkinson’s disease: advances and challenges. Lancet Neurol. 2022 Jan 1;21(1):89–102.

2. Weintraub D, Aarsland D, Biundo R, Dobkin R, Goldman J, Lewis, S. Management of psychiatric and cognitive complications in Parkinson’s disease. BMJ. 2022;e068718(379).

3. Aarsland D, Marsh L, Schrag A. Neuropsychiatric symptoms in Parkinson’s disease. Vol. 24, Movement Disorders. 2009. p. 2175–86.

4. Weintraub D, Mamikonyan E. The Neuropsychiatry of Parkinson Disease: A Perfect Storm. American Journal of Geriatric Psychiatry [Internet]. 2019;27(9):998–1018. Available from: 10.1016/j.jagp.2019.03.002

5. Braak H, Del Tredici K, Rüb U, De Vos RAI, Jansen Steur ENH, Braak E. Staging of brain pathology related to sporadic Parkinson’s disease. Neurobiol Aging. 2003 Mar 1;24(2):197–211.

6. Hawkes CH, Del Tredici K, Braak H. A timeline for Parkinson’s disease. Parkinsonism Relat Disord. 2010;16(2):79–84.

7. Schultz W. Updating dopamine reward signals. Curr Opin Neurobiol. 2013 Apr 1;23(2):229–38.

8. Cools R. Chemistry of the Adaptive Mind: Lessons from Dopamine. Neuron. 2019 Oct 9;104(1):113–31.

9. Eshel N, Roiser JP. Reward and Punishment Processing in Depression. Biol Psychiatry. 2010 Jul 15;68(2):118–24.

10. Admon R, Pizzagalli DA. Dysfunctional reward processing in depression. Curr Opin Psychol. 2015 Aug 1;4:114–8.

11. Husain M, Roiser JP. Neuroscience of apathy and anhedonia: A transdiagnostic approach. Nat Rev Neurosci. 2018;19(8):470–84.

12. Volkow ND, Wang GJ, Fowler JS, Tomasi D, Baler R. Food and drug reward: Overlapping circuits in human obesity and addiction. Curr Top Behav Neurosci [Internet]. 2011 Oct 21 [cited 2025 Sep 29];11:1–24. Available from: https://link.springer.com/chapter/10.1007/7854_2011_169

13. Le Heron C, Holroyd CB, Salamone J, Husain M. Brain mechanisms underlying apathy. J Neurol Neurosurg Psychiatry [Internet]. 2019 Mar 1 [cited 2025 Apr 28];90(3):302–12. Available from: https://jnnp.bmj.com/content/90/3/302

14. Bódi N, Kéri S, Nagy H, Moustafa A, Myers CE, Daw N, et al. Reward-learning and the novelty-seeking personality: A between-and within-subjects study of the effects of dopamine agonists on young parkinsons patients. Brain. 2009;132(9):2385–95.

15. Rutledge RB, Lazzaro SC, Lau B, Myers CE, Gluck MA, Glimcher PW. Dopaminergic drugs modulate learning rates and perseveration in Parkinson’s patients in a dynamic foraging task. Journal of Neuroscience. 2009 Dec 2;29(48):15104–14.

16. McCoy B, Jahfari S, Engels G, Knapen T, Theeuwes J. Dopaminergic medication reduces striatal sensitivity to negative outcomes in Parkinson’s disease. Brain. 2019;142(11):3605–20.

17. Sun S, Madge V, Djordjevic J, Gagnon JF, Collins DL, Dagher A, et al. Selective Effects of Substantia Nigra and Locus Coeruleus Degeneration on Cognition in Parkinson’s Disease. Movement Disorders. 2025 May 1;40(5):844–54.

18. Timmer MHM, Sescousse G, Van Der Schaaf ME, Esselink RAJ, Cools R. Reward learning deficits in Parkinson’s disease depend on depression. Psychol Med [Internet]. 2017 Oct 1 [cited 2022 Jun 5];47(13):2302–11. Available from: https://www.cambridge.org/core/journals/psychological-medicine/article/reward-learning-deficits-in-parkinsons-disease-depend-on-depression/6487DD029FD65BB9DF2F162CD0C0E57A

19. Herzallah MM, Khdour HY, Taha AB, Elmashala AM, Mousa HN, Taha MB, et al. Depression reduces accuracy while parkinsonism slows response time for processing positive feedback in patients with Parkinson’s disease with comorbid major depressive disorder tested on a probabilistic category-learning task. Front Psychiatry. 2017 Jun 12;8(JUN).

20. Costello H, Yamamori Y, Kieslich K, Murphy M, Bobyreva K, Schrag AE, et al. Impaired reward sensitivity in Parkinson’s depression is unresponsive to dopamine treatment. Brain [Internet]. 2025 Jun 3 [cited 2025 Sep 29];148(6):2122–34. Available from: 10.1093/brain/awaf098

21. Buelow MT, Frakey LL, Grace J, Friedman JH. The contribution of apathy and increased learning trials to risky decision-making in Parkinson’s disease. Archives of Clinical Neuropsychology. 2014;29(1):100–9.

22. Gilmour W, Mackenzie G, Feile M, Tayler-Grint L, Suveges S, Macfarlane JA, et al. Impaired value-based decision-making in Parkinson’s disease apathy. Brain [Internet]. 2024 Apr 4 [cited 2025 Jan 19];147(4):1362–76. Available from: 10.1093/brain/awae025

23. Voon V, Pessiglione M, Brezing C, Gallea C, Fernandez HH, Dolan RJ, et al. Mechanisms Underlying Dopamine-Mediated Reward Bias in Compulsive Behaviors. Neuron. 2010 Jan 14;65(1):135–42.

24. Piray P, Zeighami Y, Bahrami F, Eissa AM, Hewedi DH, Moustafa AA. Impulse control disorders in Parkinson’s disease are associated with dysfunction in stimulus valuation but not action valuation. Journal of Neuroscience. 2014;34(23):7814–24.

25. Costello H, Berry AJ, Reeves S, Weil RS, Joyce EM, Howard R, et al. Disrupted reward processing in Parkinson’s disease and its relationship with dopamine state and neuropsychiatric syndromes: a systematic review and meta-analysis. J Neurol Neurosurg Psychiatry. 2022;93(5):555–62.

26. Insel T, Cuthbert B, Garvey M, Heinssen R, Pine DS, Quinn K, et al. Research Domain Criteria (RDoC): Toward a New Classification Framework for Research on Mental Disorders. 101176/appi.ajp201009091379 [Internet]. 2010 Jul [cited 2025 Jan 21];167(7):748–51. Available from: https://psychiatryonline.org/doi/10.1176/appi.ajp.2010.09091379

27. Frank MJ, Seeberger LC, O’Reilly RC. By carrot or by stick: Cognitive reinforcement learning in Parkinsonism. Science (1979). 2004;306(5703):1940–3.

28. Pizzagalli DA, Jahn AL, O’Shea JP. Toward an Objective Characterization of an Anhedonic Phenotype: A Signal-Detection Approach. Biol Psychiatry. 2005;57(4):319–27.

29. Frank MJ, Samanta J, Moustafa AA, Sherman SJ. Hold Your Horses: Impulsivity, Deep Brain Stimulation, and Medication in Parkinsonism. Science (1979) [Internet]. 2007;318(November):1309–12. Available from: http://www.ncbi.nlm.nih.gov/pubmed/17962524

30. Shiner T, Seymour B, Wunderlich K, Hill C, Bhatia KP, Dayan P, et al. Dopamine and performance in a reinforcement learning task: evidence from Parkinson’s disease. Brain [Internet]. 2012 Jun 1 [cited 2022 Jun 5];135(6):1871–83. Available from: https://academic.oup.com/brain/article/135/6/1871/329665

31. Pizzagalli DA, Evins AE, Schetter EC, Frank MJ, Pajtas PE, Santesso DL, et al. Single Dose of a Dopmaine Agonist Impairs Reinforcement Learning in Humans: Behavioral Evidence from a Laboratory-based Measure of Reward Responsiveness. Psychopharmacology (Berl). 2008;196(2):221–32.

32. Vrieze E, Ceccarini J, Pizzagalli DA, Bormans G, Vandenbulcke M, Demyttenaere K, et al. Measuring extrastriatal dopamine release during a reward learning task. Hum Brain Mapp. 2013;34(3):575–86.

33. Pizzagalli D, Iosifescu D, Hallett L, Ratner K, Maurizo F. Reduced Hedonic Capacity in Major Depressive Disorder: Evidence from a Probabilistic Reward Task. J Psychiatr Res. 2009;43(1):76–87.

34. Gan-Or Z, Rao T, Leveille E, Degroot C, Chouinard S, Cicchetti F, et al. The Quebec Parkinson Network: A Researcher-Patient Matching Platform and Multimodal Biorepository. J Parkinsons Dis [Internet]. 2020 [cited 2024 Oct 7];10(1):301–13. Available from: /pmc/articles/PMC7029361/

35. Sharp ME, Duncan K, Foerde K, Shohamy D. Dopamine is associated with prioritization of reward-associated memories in Parkinson’s disease. Brain. 2020;143(8):2519–31.

36. Pizzagalli DA, Goetz E, Ostacher M, Iosifescu D V., Perlis RH. Euthymic Patients with Bipolar Disorder Show Decreased Reward Learning in a Probabilistic Reward Task. Biol Psychiatry [Internet]. 2008 Jul 15 [cited 2024 Nov 6];64(2):162. Available from: https://pmc.ncbi.nlm.nih.gov/articles/PMC2464620/

37. Vrieze E, Pizzagalli DA, Demyttenaere K, Hompes T, Sienaert P, De Boer P, et al. Reduced Reward Learning Predicts Outcome in Major Depressive Disorder. Biol Psychiatry. 2013 Apr 1;73(7):639–45.

38. Whitton AE, Kumar P, Treadway MT, Rutherford A V., Ironside ML, Foti D, et al. Mapping Disease Course Across the Mood Disorder Spectrum Through a Research Domain Criteria Framework. Biol Psychiatry Cogn Neurosci Neuroimaging [Internet]. 2021 Jul 1 [cited 2021 Jul 18];6(7):706–15. Available from: http://www.biologicalpsychiatrycnni.org/article/S2451902221000239/fulltext

39. Yesavage JA, Brink TL, Rose TL, Lum O, Huang V, Adey M, et al. Development and validation of a geriatric depression screening scale: A preliminary report. J Psychiatr Res. 1982 Jan 1;17(1):37–49.

40. Lesher EL, Berryhill JS. Validation of the geriatric depression scale short form among inpatients. J Clin Psychol. 1994;50(2):256–60.

41. Herrmann N, Bust0 UA, Mittmann N, Silver IL, Shulman KI, Shear Claud10 NH, et al. A VALIDATION STUDY OF THE GERIATRIC DEPRESSION SCALE SHORT FORM. Int J Geriatr Psychiatry [Internet]. 1996 [cited 2025 Jan 21];11:457–60. Available from: https://onlinelibrary.wiley.com/doi/10.1002/

42. Snaith RP, Hamilton M, Morley S, Humayan A, Hargreaves D, Trigwell P. A scale for the assessment of hedonic tone. The Snaith-Hamilton Pleasure Scale. British Journal of Psychiatry. 1995;167(JULY):99–103.

43. Marin RS, Biedrzycki RC, Firinciogullari S. Reliability and validity of the apathy evaluation scale. Psychiatry Res. 1991 Aug 1;38(2):143–62.

44. Patton JH, Stanford MS, Barratt ES. FACTOR STRUCTURE OF THE BARRATT IMPULSIVENESS SCALE. [cited 2025 Jan 21]; Available from: https://onlinelibrary.wiley.com/terms-and-conditions

45. Weintraub D, Hoops S, Shea JA, Lyons KE, Pahwa R, Driver-Dunckley ED, et al. Validation of the questionnaire for impulsive-compulsive disorders in Parkinson’s disease. Movement Disorders [Internet]. 2009 Jul 30 [cited 2025 Jan 21];24(10):1461–7. Available from: https://onlinelibrary.wiley.com/doi/full/10.1002/mds.22571

46. Muhammed K, Manohar S, Ben Yehuda M, Chong TTJ, Tofaris G, Lennox G, et al. Reward sensitivity deficits modulated by dopamine are associated with apathy in Parkinson’s disease. Brain. 2016 Oct 1;139(10):2706–21.

47. Martínez-Horta S, Riba J, de Bobadilla RF, Pagonabarraga J, Pascual-Sedano B, Antonijoan RM, et al. Apathy in Parkinson’s Disease: Neurophysiological Evidence of Impaired Incentive Processing. The Journal of Neuroscience [Internet]. 2014 [cited 2025 Oct 5];34(17):5918. Available from: https://pmc.ncbi.nlm.nih.gov/articles/PMC6608288/

48. Lueken U, Evens R, Balzer-Geldsetzer M, Baudrexel S, Dodel R, Gräber-Sultan S, et al. Psychometric properties of the apathy evaluation scale in patients with Parkinson’s disease. Int J Methods Psychiatr Res [Internet]. 2017 Dec 1 [cited 2024 Aug 4];26(4):e1564. Available from: https://onlinelibrary.wiley.com/doi/full/10.1002/mpr.1564

49. Voon V, Sohr M, Lang AE, Potenza MN, Siderowf AD, Whetteckey J, et al. Impulse control disorders in parkinson disease: A multicenter case–control study. Ann Neurol [Internet]. 2011 Jun 1 [cited 2025 Apr 27];69(6):986–96. Available from: 10.1002/ana.22356

50. Housden CR, O’Sullivan SS, Joyce EM, Lees AJ, Roiser JP. Intact reward learning but elevated delay discounting in Parkinson’s disease patients with impulsive-compulsive spectrum behaviors. Neuropsychopharmacology. 2010 Oct;35(11):2155–64.

51. Drew DS, Muhammed K, Baig F, Kelly M, Saleh Y, Sarangmat N, et al. Dopamine and reward hypersensitivity in Parkinson’s disease with impulse control disorder. Brain. 2020 Aug 1;143(8):2502–18.

52. Barber TR, Griffanti L, Muhammed K, Drew DS, Bradley KM, McGowan DR, et al. Apathy in rapid eye movement sleep behaviour disorder is associated with serotonin depletion in the dorsal raphe nucleus. Brain [Internet]. 2018 Oct 1 [cited 2024 Dec 2];141(10):2848–54. Available from: 10.1093/brain/awy240

53. McGuigan S, Zhou SH, Brosnan MB, Thyagarajan D, Bellgrove MA, Chong TTJ. Dopamine restores cognitive motivation in Parkinson’s disease. Brain. 2019 Mar 1;142(3):719–32.

54. Le Heron C, Plant O, Manohar S, Ang YS, Jackson M, Lennox G, et al. Distinct effects of apathy and dopamine on effort-based decision-making in Parkinson’s disease. Brain [Internet]. 2018 May 1 [cited 2021 Nov 10];141(5):1455–69. Available from: https://academic.oup.com/brain/article/141/5/1455/4974326

55. van der Velden RMJ, Broen MPG, Kuijf ML, Leentjens AFG. Frequency of mood and anxiety fluctuations in Parkinson’s disease patients with motor fluctuations: A systematic review. Movement Disorders [Internet]. 2018 Oct 1 [cited 2025 Dec 24];33(10):1521–7. Available from: 10.1002/mds.27465

56. van der Velden RMJ, Mulders AEP, Drukker M, Kuijf ML, Leentjens AFG. Network analysis of symptoms in a Parkinson patient using experience sampling data: An n = 1 study. Movement Disorders [Internet]. 2018 Dec 1 [cited 2025 Dec 24];33(12):1938–44. Available from: 10.1002/mds.93

57. Brown VM, Chen J, Gillan CM, Price RB. Improving the reliability of computational analyses: Model-based planning and its relationship with compulsivity. Biol Psychiatry Cogn Neurosci Neuroimaging [Internet]. 2020 Jun 1 [cited 2025 Dec 24];5(6):601. Available from: https://pmc.ncbi.nlm.nih.gov/articles/PMC7286766/

58. Schaaf J V., Weidinger L, Molleman L, van den Bos W. Test–retest reliability of reinforcement learning parameters. Behavior Research Methods 2023 56:5 [Internet]. 2023 Sep 8 [cited 2025 Dec 24];56(5):4582–99. Available from: https://link.springer.com/article/10.3758/s13428-023-02203-4

59. Vrizzi S, Najar A, Lemogne C, Palminteri S, Lebreton M. Behavioral, computational and self-reported measures of reward and punishment sensitivity as predictors of mental health characteristics. Nature Mental Health. 2025 Jun 1;3(6):654–66.

60. Zeynep Enkavi A, Eisenberg IW, Bissett PG, Mazza GL, MacKinnon DP, Marsch LA, et al. Large-scale analysis of test–retest reliabilities of self-regulation measures. Proc Natl Acad Sci U S A. 2019;116(12):5472–7.

61. Enkavi AZ, Poldrack RA. Implications of the Lacking Relationship Between Cognitive Task and Self-report Measures for Psychiatry. Biol Psychiatry Cogn Neurosci Neuroimaging [Internet]. 2021;6(7):670–2. Available from: 10.1016/j.bpsc.2020.06.010

62. Maillet A, Météreau E, Tremblay L, Favre E, Klinger H, Lhommée E, et al. Serotonergic and Dopaminergic Lesions Underlying Parkinsonian Neuropsychiatric Signs. Movement Disorders [Internet]. 2021 Dec 1 [cited 2025 Jan 21];36(12):2888–900. Available from: https://onlinelibrary.wiley.com/doi/full/10.1002/mds.28722

63. Mayeux R, Stern Y, Sano M, Williams JBW, Cote LJ. The relationship of serotonin to depression in Parkinson’s disease. Movement Disorders. 1988;3(3):237–44.

64. Marsh L, Biglan K, Gerstenhaber M, Williams JR. Atomoxetine for the treatment of executive dysfunction in Parkinson’s disease: A pilot open-label study. Mov Disord [Internet]. 2009 Jan 1 [cited 2024 Apr 10];24(2):277–82. Available from: /pmc/articles/PMC2683743/

65. Prasuhn J, Prasuhn M, Fellbrich A, Strautz R, Lemmer F, Dreischmeier S, et al. Association of Locus Coeruleus and Substantia Nigra Pathology With Cognitive and Motor Functions in Patients With Parkinson Disease. Neurology. 2021;97(10):e1007–16.

66. O’Callaghan C, Lewis SJG. Cognition in Parkinson’s Disease [Internet]. 1st ed. Vol. 133, International Review of Neurobiology. Elsevier Inc.; 2017. 557–583 p. Available from: 10.1016/bs.irn.2017.05.002

