## Supplementary Materials for "Exploring reward learning disruptions as a possible mechanism underlying neuropsychiatric symptoms in Parkinson’s disease"

**Methods**

**Reward Learning Tasks**

*Probabilistic Stimulus Selection Task: Test Phase.* In the test phase of the task, participants were shown the same three training pairs (i.e., AB, CD, EF) and eight new combinations of the stimuli (**Supplementary** **Fig. 1B**) and were asked to select the stimulus associated with greater probability of reward. These new combinations were constructed such that the stimulus associated with the greatest probability of reward was paired with all other stimuli (i.e., AC, AD, AE, AF) the stimulus associated with the lowest probability of reward was paired with all other stimuli (i.e., BC, BD, BE, BF). For each of these new pairings, if participants successfully learned about the reward values of each stimulus, they should select stimulus A (approach A) or avoid selecting stimulus B (avoid B). Each pair was presented 10 times in random order (a total of 110 trials).

*Probabilistic Reward Task.* On each trial, participants were presented with a cartoon face. Either a short (11.5 mm) or long (13 mm) mouth would briefly appear for 100 ms. Participants had to identify which stimulus length was presented using the keyboard (“z” for short and “m” for long). Across all trials, participants were shown an equal number of short and long stimuli and received intermittent feedback for correct responses only in the form of “50 points” onscreen (**Supplementary** **Fig. 1C**). The stimulus associated with a leaner reward schedule resulted in rewarding feedback for 20 percent of correct responses, while the stimulus associated with a richer reward schedule resulted in rewarding feedback for 60 percent of correct responses. Whether the short or long mouth was assigned the richer reward schedule was counterbalanced across participants. The task consisted of three blocks of 100 trials with equal presentations of both stimulus types within each block. We used the task parameters described in Pizzagalli et al., 2005.

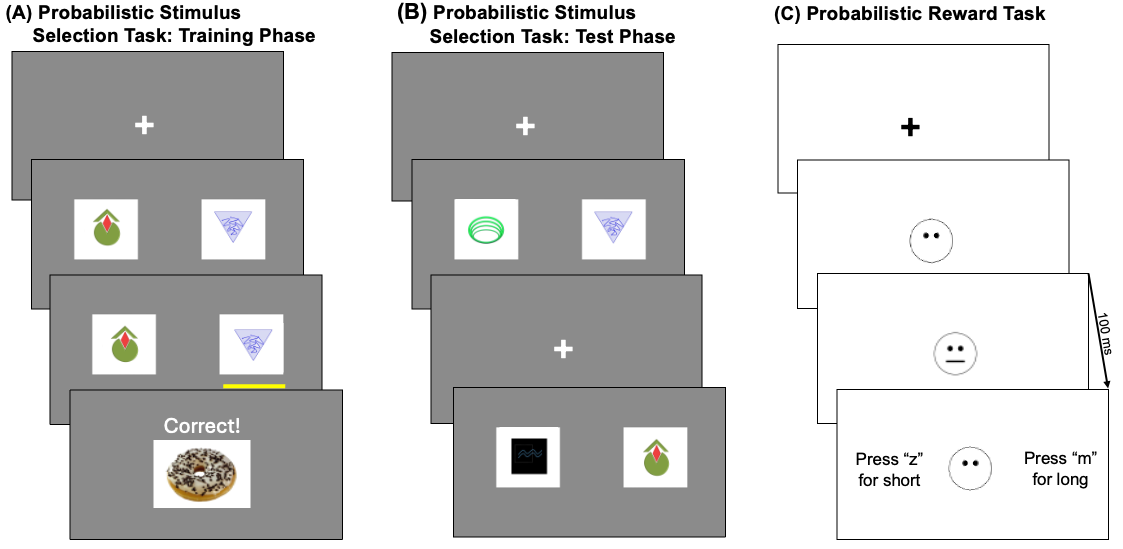

**Supplementary Figure 1 Reward Learning Tasks. (A)** In the training phase of the probabilistic stimulus selection task, participants are tasked with learning the reward values of three pairs of stimuli (AB, CD, EF) through feedback across 150 trials. One example trial is depicted. **(B)** In the test phase of the probabilistic stimulus selection task, participants must select which stimulus they think is associated with greater chance of reward based on what they learned in the training phase. The training pairings are shown along with new pairings where the most and least likely rewarded stimuli are paired with all other stimuli (AC, AD, AE, AF, BC, BD, BE, BF). No feedback is given. Two example trials are depicted. **(C)** In the Probabilistic Reward Task, despite equal presentations of the short (11.5 mm) and long (13 mm) mouths, one mouth is rewarded three times more often to induce a response bias for the richly rewarded mouth. One example trial with no reward is depicted.

**Results**

**Reward learning summary measures**

*Positive learning rate.* Computationally derived positive learning rates have been previously associated with substantia nigra degeneration, as measured by neuromelanin signal intensity, and with dopamine state in PD patients (Rutledge et al., 2009; Sun et al., 2025). We derived positive learning rates by applying a reinforcement learning drift diffusion model to the training phase data from the probabilistic stimulus selection task (previously described in detail in Sun et a., 2025). The model uses Bayesian and hierarchical estimation. As such, we evaluated discovery and replication sample differences by computing the posterior probability, i.e., the probability that the posterior distributions of the positive learning rates for both groups are the same. Positive learning rates represent the ability to adapt responding based on positive feedback.

*Learning Slope.* Mixed effect models account for within- and between-participant variability (Hoffman & Rovine 2007) and have been previously applied to reward learning task performance in PD (Sharp et al., 2016; Sharp et al., 2020). We derived learning slopes from a logistic mixed effects model of the training phase data from the probabilistic stimulus selection task. The dependent variable was ‘accuracy coded as 0 if the participant did not select the stimulus associated with greater reward probability and 1 if the participant selected the stimulus associated with greater reward probability. We grouped trials into 10 blocks of 15 trials and included ‘block’ (1-10) and ‘sample’ (Samples 1 and 2 effect coded as -0.5 and 0.5 respectively) as fixed effects. We interacted ‘block’ and ‘sample’ to test whether there were sample differences in the learning slopes. We also included a random intercept and ‘block’ as a random slope (1).

(1) Response ~ 1 + block + sample + block × sample + (1 + block | participant)

Learning slopes for each participant were calculated as the sum of the fixed and random effects for block such that the fixed effect represents the relationship between ‘block’ and ‘response’ on average across participants and the random effect represents the individual deviation from the average for each participant.

*Approach ‘A’ Accuracy.* Performance in the test phase of the probabilistic stimulus selection has been previously evaluated as approach ‘A’ and avoid ‘B’ accuracy representing the respective accuracies in selecting the stimulus associated with the highest probability of yielding a reward and avoiding the stimulus associated with the lowest probability of yielding a reward. In the present study, we focused on the approach ‘A’ accuracy as this represents the ability to learn from positive feedback and because previous PD studies have demonstrated that approach ‘A’ performance is better than avoid ‘B’ performance when in the dopamine medicated state (Frank et al., 2004; Frank et al., 2007; Shiner et al., 2012). We computed approach ‘A’ accuracy for each participant by averaging the responses on test trials for stimulus ‘A’, i.e., the stimulus that is associated with the highest probability of reward (trials: AC, AD, AE, AF). Sample differences were evaluated using an independent samples t-test on approach ‘A’ accuracy.

*Response bias.* For this outcome measure, we used performance on the probabilistic reward task. Previous research utilizing this task have summarized performance using a response bias, which describes the magnitude of biased responding for the stimulus that is rewarded more often. Studies have shown that individuals with depression have reduced response biases compared to controls (Pizzagalli et al., 2005; Pizzagalli et al., 2008; Vrieze et al., 2013). In keeping with this body of work, we derived response bias for each participant from performance on the probabilistic reward task as (2):

(2) $\log b=0.5\times log(\frac{{Rich}_{Correct} \times{Lean}_{Incorrect}}{{Rich}_{Incorrect} \times{Rich}_{Correct}})$

The trials were divided into three blocks of 100 trials each and Δ response bias was calculated by subtracting the response bias in block 1 from the response bias in block 3 such that a greater Δ response bias indicates greater learning of the response bias. Sample differences in Δ response bias was evaluated using an independent samples t-test.

*Rich-Lean Hit Rate.* For this outcome measure, we used performance on the probabilistic reward task. Previous work using this measure showed that individuals with depression have a smaller discrepancy between rich and lean hit rates compared to controls (Pizzagalli et al., 2009). The present study reproduced this measure by calculating the difference in hit rates between rich and lean stimuli. Sample differences in rich-lean hit rate was evaluated using an independent samples t-test.

**Results**

*Sample differences in reward learning measures.* RLDDM-derived positive learning rates from the training phase of the probabilistic stimulus selection task did not differ across samples (Mean_Sample1_ = 0.125, Mean_Sample2_ = 0.130, *p* = 0.556). One participant in each sample was removed from further analyses due to lack of parameter convergence.

The logistic mixed-effects model of performance in the training phase of the probabilistic stimulus selection task revealed that across both samples, the odds of selecting the stimulus associated with greater reward increased by 7 percent for each block (*β_block_* = 0.07, *p* < 0.0001), indicating that there was learning overall. On average, there was no difference in odds of selecting the stimulus associated with greater reward between samples (*β_sample_* = 0.16, *p* = 0.22) and no interaction between sample and block (*β_block*sample_* = 0.006, *p* = 0.80).

Approach ‘A’ accuracy from the test phase of the probabilistic stimulus selection task did not differ across samples (*t*_(166)_ = -0.70, *p* = 0.49). Three participants in the discovery sample did not complete the test phase of the reinforcement learning task due to fatigue.

The Δ response bias and the difference in rich-lean hit rate from the probabilistic reward task did not differ across samples (*t*_(137)_ = 0.43, *p* = 0.67). Participants were excluded from analyses because of task programming errors, incomplete data, or incorrectly responding on all trials so only 58 and 81 participants from the samples 1 and 2 respectively were included in the analysis (**Supplementary Table 1**).

In summary, the samples did not differ in performance across all measures of reward learning.

**Sample 1	Sample 2**

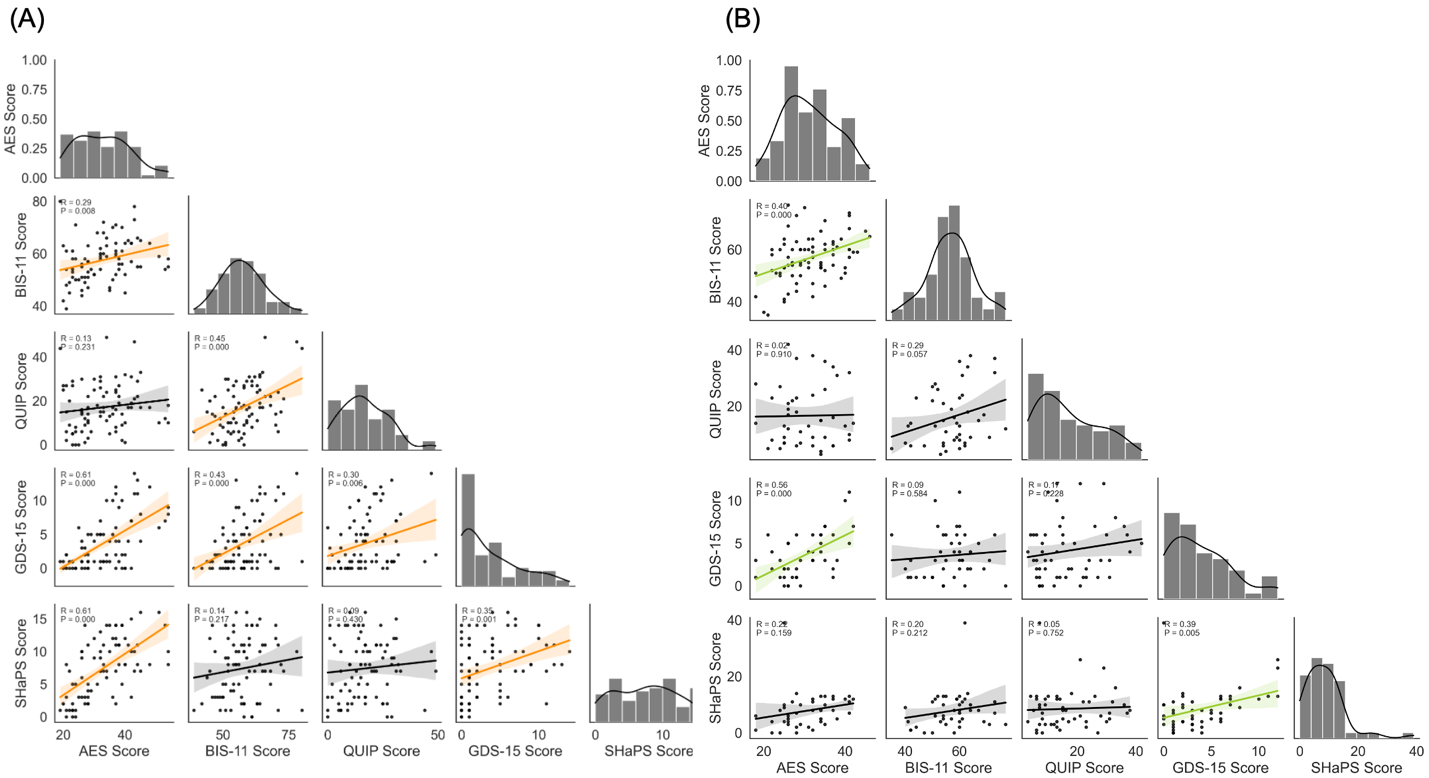

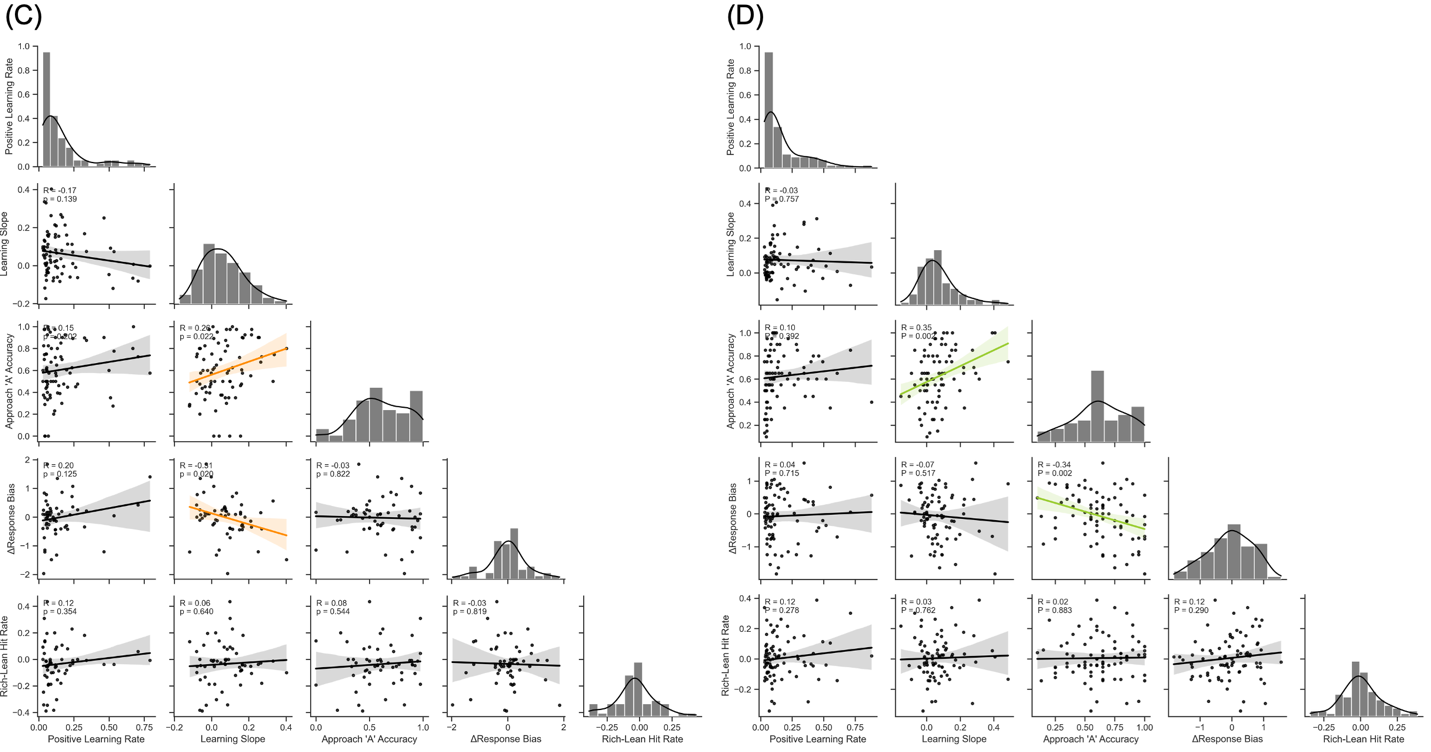

**Supplementary Figure 2.** Correlation matrix of (A) sample 1 neuropsychiatric symptom scores, (B) sample 2 neuropsychiatric symptom scores, (C) sample 1 reward learning summary measures, and (D) sample 2 reward learning summary measures. Coloured regression lines depict significant correlations. Histograms for each variable are shown along the diagonal. Abbreviations: Apathy Evaluation Scale (AES), Barratt Impulsiveness Scale version 11 (BIS-11), Questionnaire for Impulsive-Compulsive Disorders in Parkinson’s disease (QUIP), Geriatric Depression Scale short form (GDS-15), Snaith Hamilton Pleasure Scale (SHaPS).

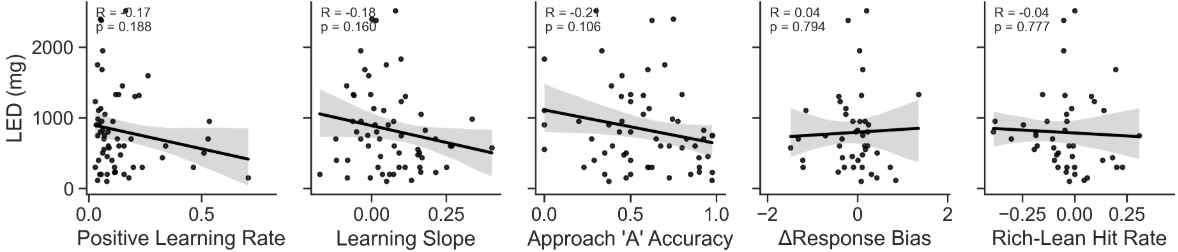

**Supplementary Figure 3.** No significant correlations between reward learning summary measures and levodopa equivalent dose (LED) in the discovery sample.

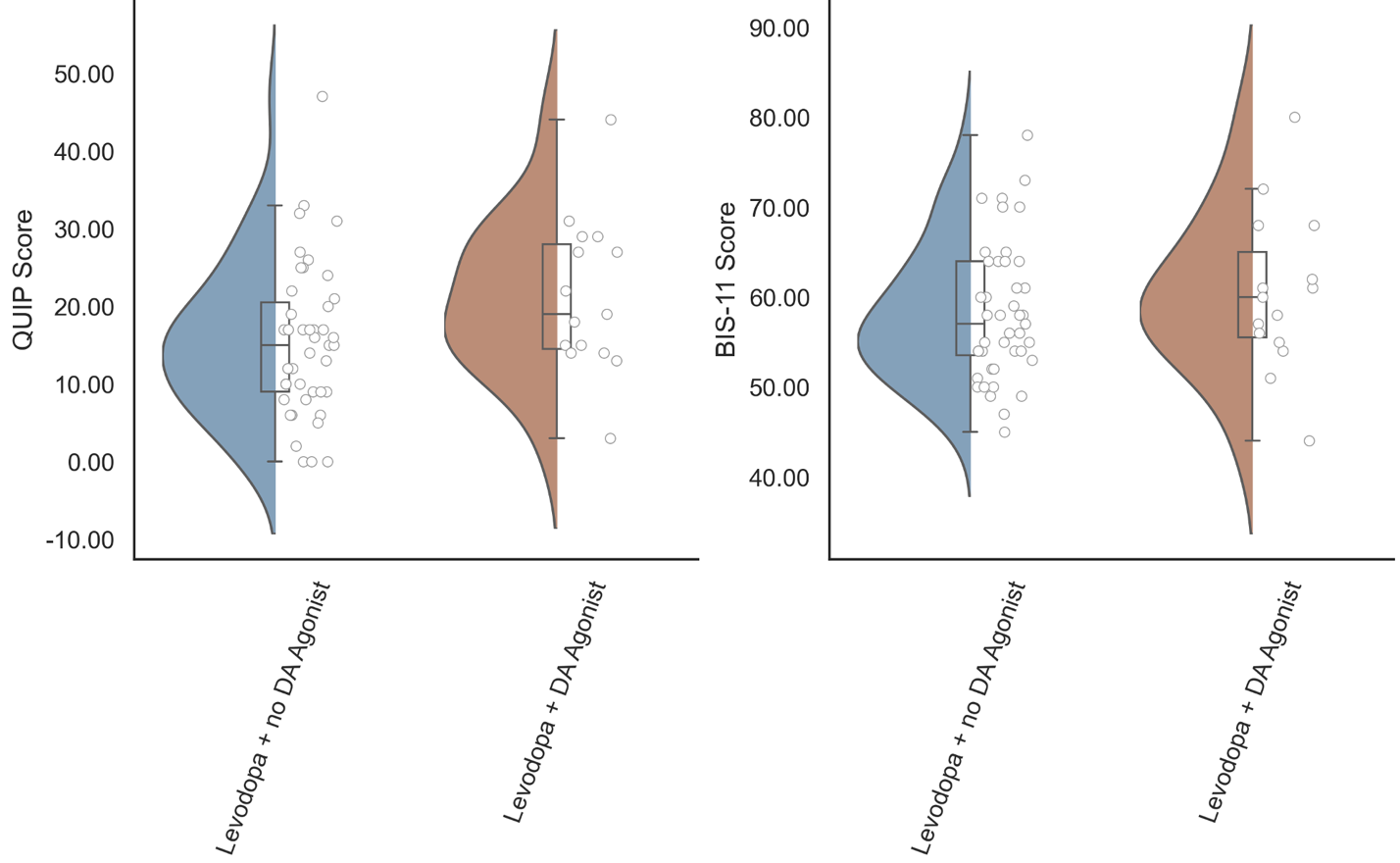

**Supplementary Figure 4.** Impulsivity scores (QUIP and BIS-11 scores) do not differ across medication regimens in sample 1.

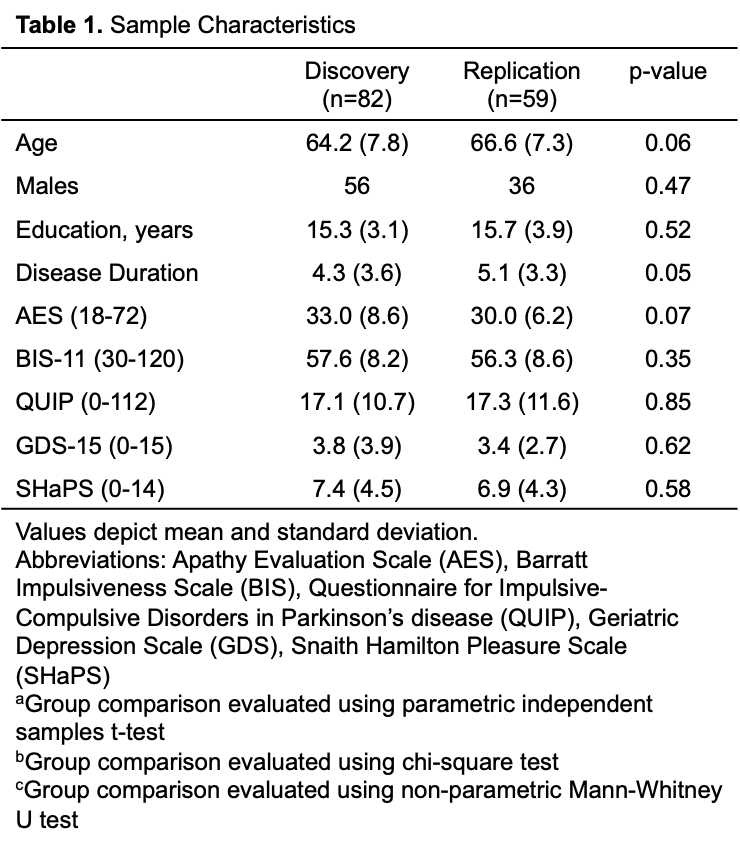

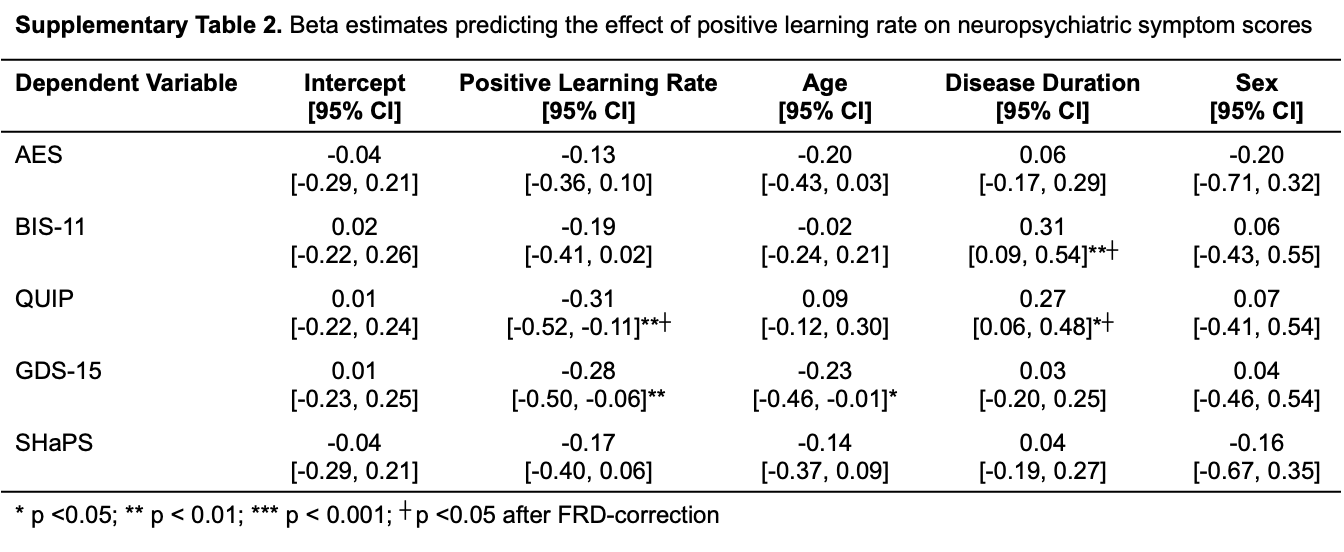

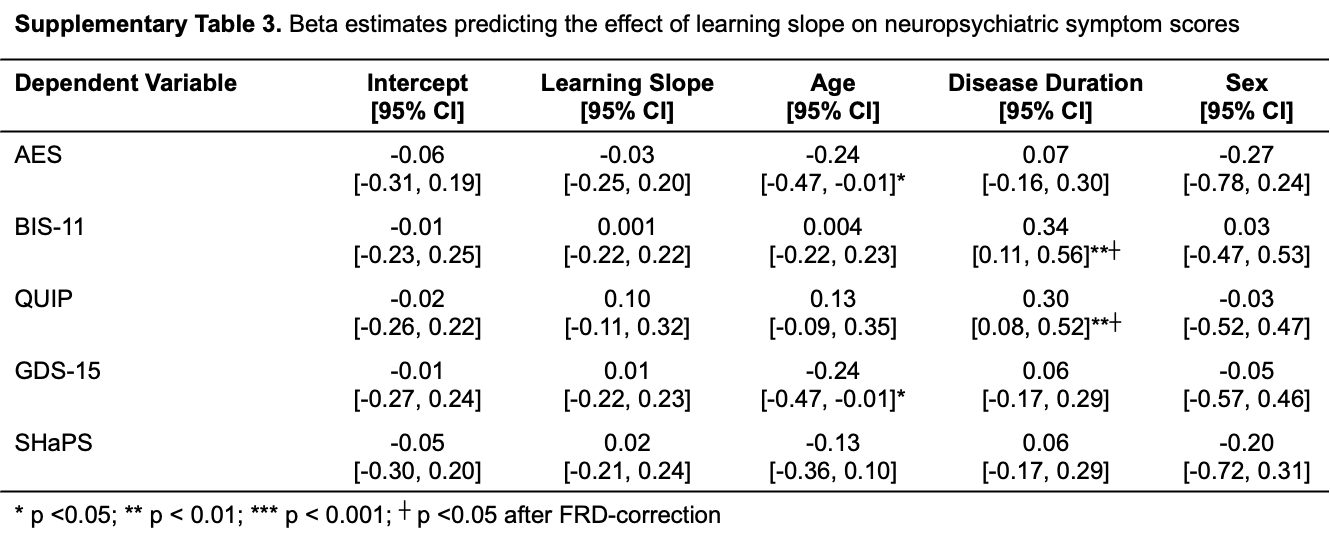

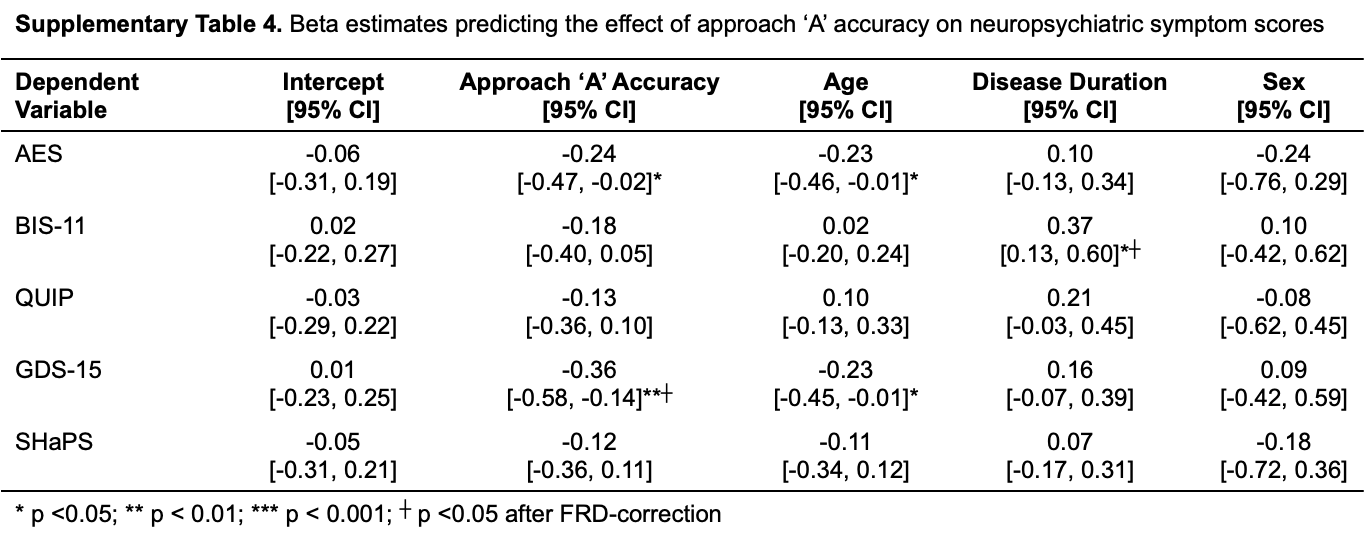

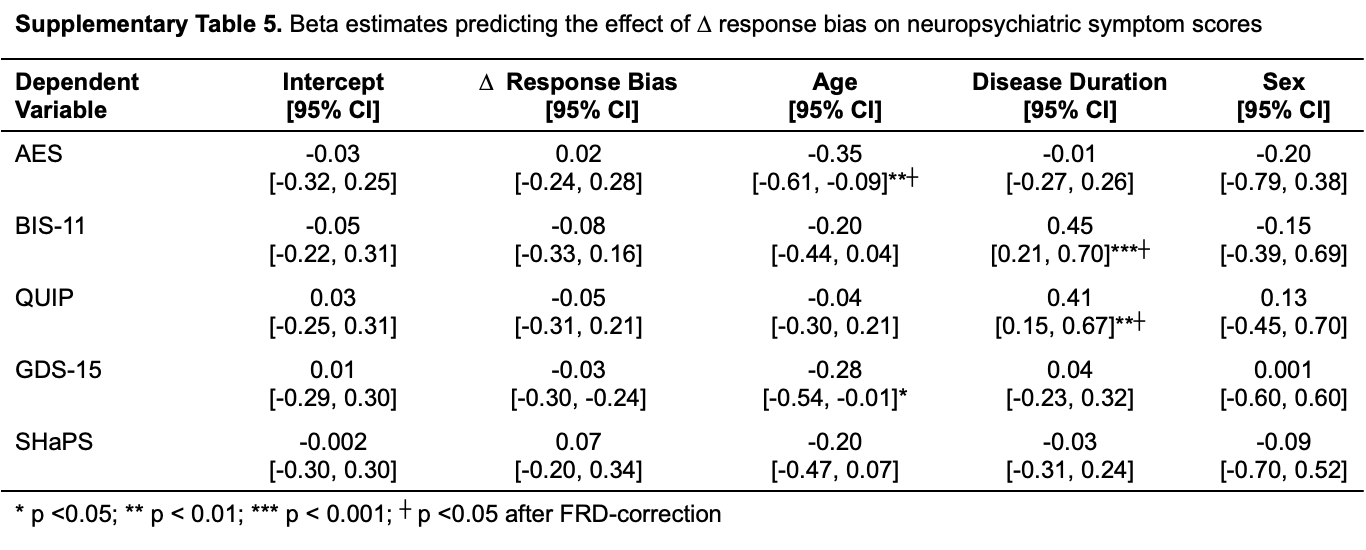

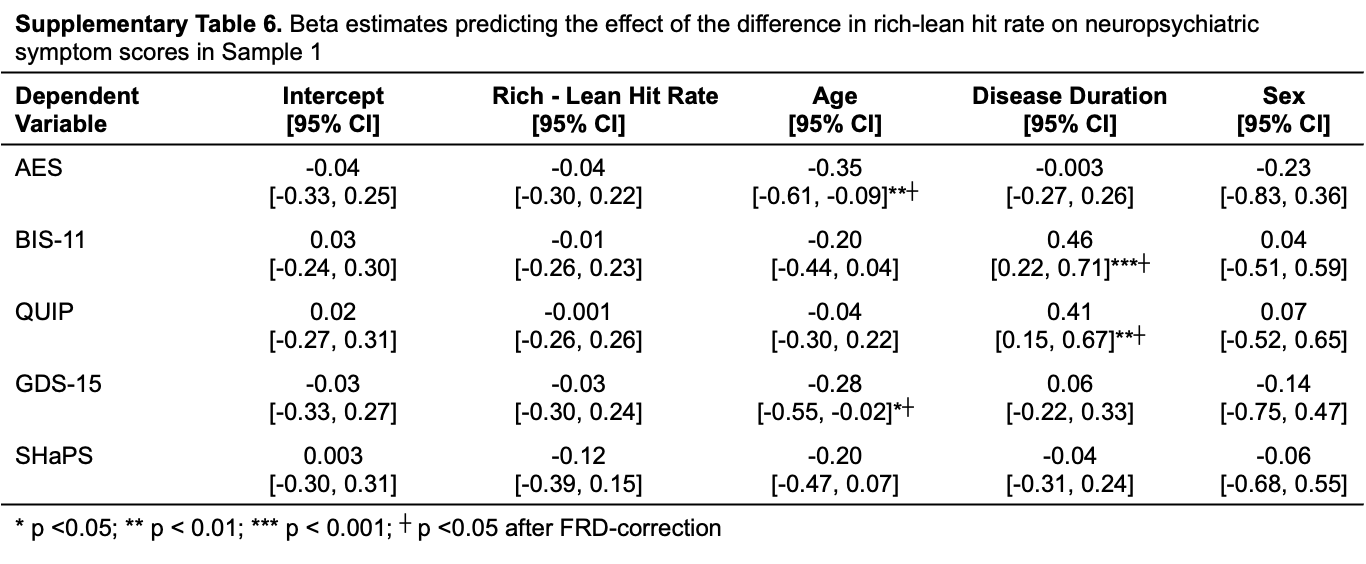

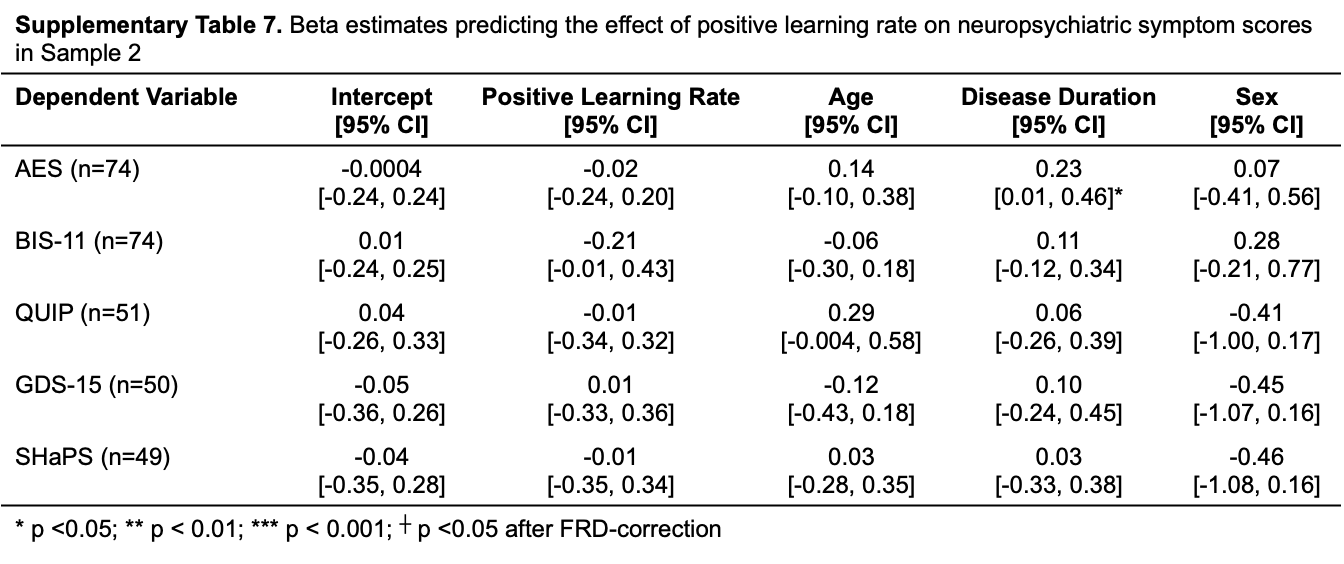

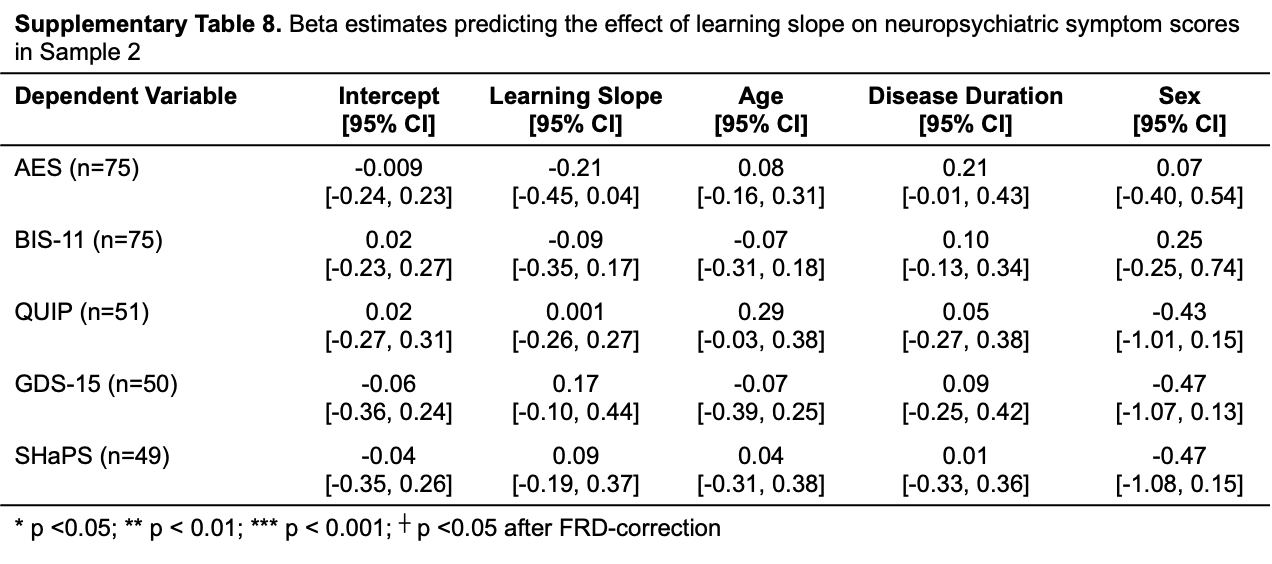

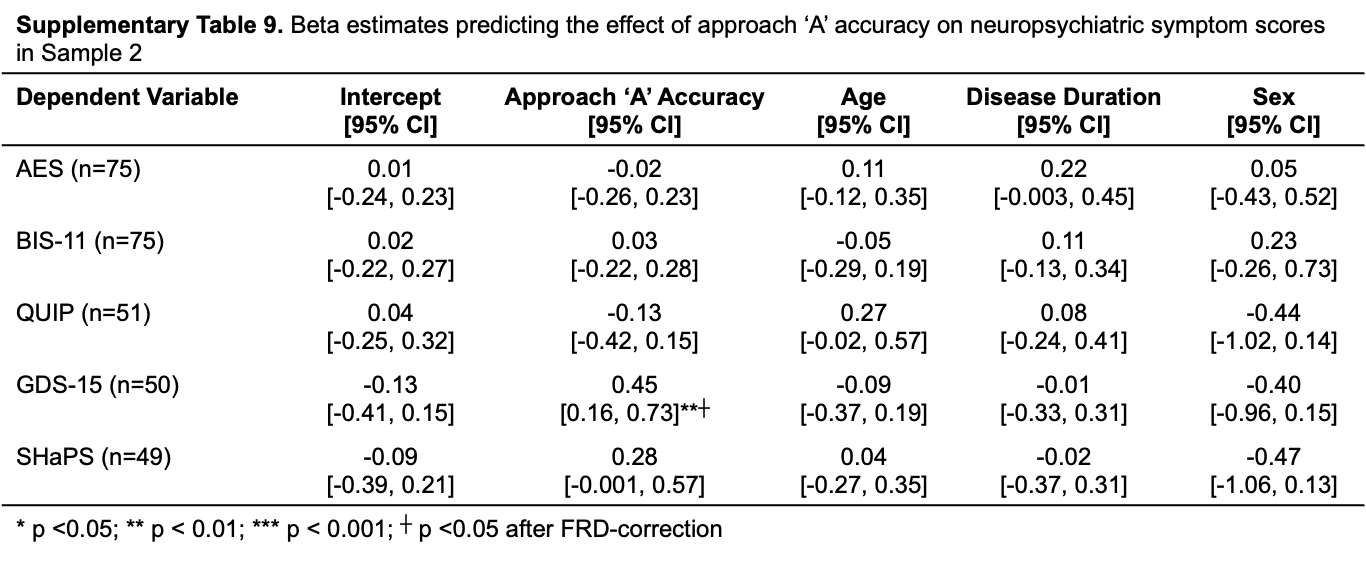

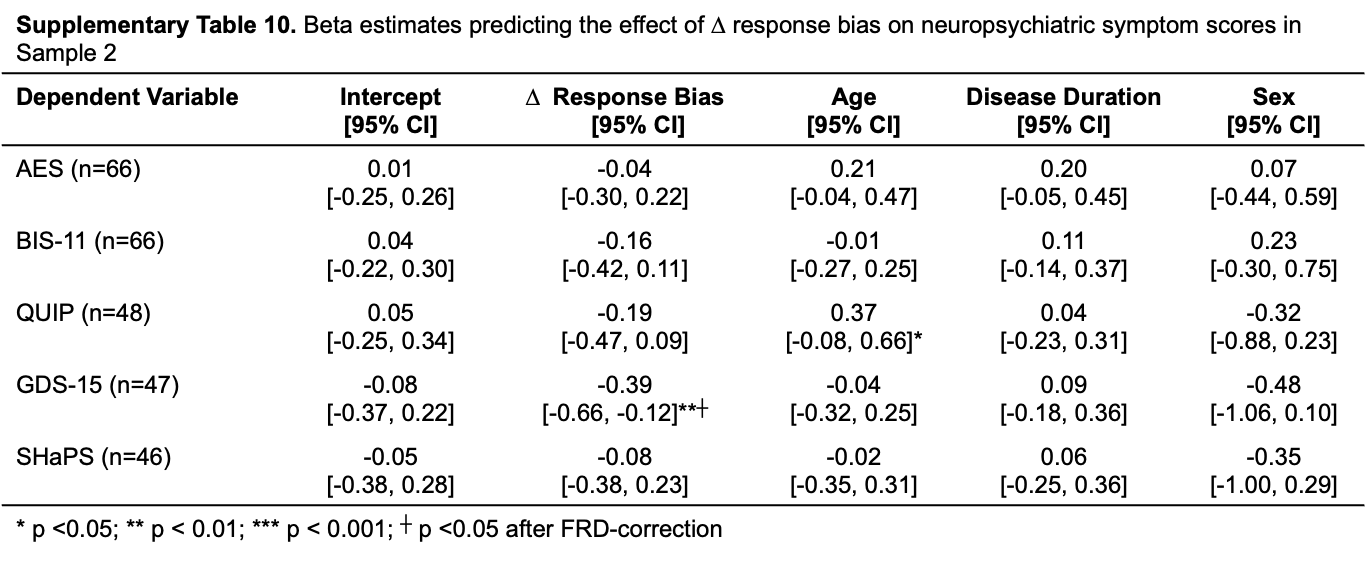

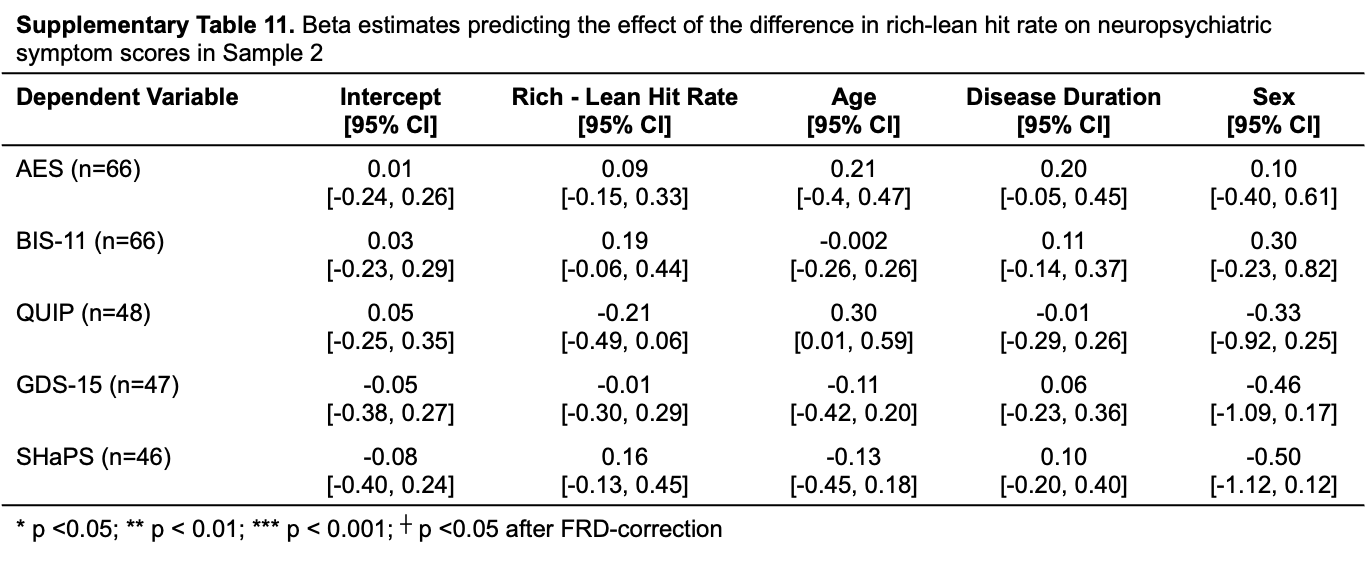
